# Emerging models and trends in mental health crisis care in England: a national investigation of crisis care systems

**DOI:** 10.1101/2021.07.08.21259617

**Authors:** Christian Dalton-Locke, Sonia Johnson, Jasmine Harju-Seppänen, Natasha Lyons, Luke Sheridan Rains, Ruth Stuart, Amelia Campbell, Jeremy Clark, Aisling Clifford, Laura Courtney, Ceri Dare, Kelly Kathleen, Chris Lynch, Paul McCrone, Shilpa Nairi, Karen Newbigging, Patrick Nyikavaranda, David Osborn, Karen Persaud, Martin Stefan, Brynmor Lloyd-Evans

## Abstract

**Background:** Inpatient psychiatric care is unpopular and expensive, and development and evaluation of alternatives is a long-standing policy and research priority around the world. In England, the three main models documented over the past fifty years (teams offering crisis assessment and treatment at home; acute day services; and residential crisis services in the community) have recently been augmented by several new service models. These are intended to enhance choice and flexibility within catchment area acute care systems, but remain largely undocumented in the research literature. We therefore aimed to describe the types and distribution of crisis care models across England through a national survey.

**Methods:** We carried out comprehensive mapping of crisis resolution teams (CRTs) using previous surveys, websites and multiple official data sources. Managers of CRTs were invited to participate as key informants who were familiar with the provision and organisation of crisis care services within their catchment area. The survey could be completed online or via telephone interview with a researcher, and elicited details about types of crisis care delivered in the local catchment area.

**Results:** We mapped a total of 200 adult CRTs and completed the survey with 184 (92%). Of the 200 mapped adult CRTs, there was a local (i.e., within the adult CRT catchment area) children and young persons CRT for 84 (42%), and an older adults CRT for 73 (37%). While all but one health region in England provided CRTs for working age adults, there was high variability regarding provision of all other community crisis service models and system configurations. Crisis cafes, street triage teams and separate crisis assessment services have all proliferated since a similar survey in 2016, while provision of acute day units has reduced.

**Conclusions:** The composition of catchment area crisis systems varies greatly across England and popularity of models seems unrelated to strength of evidence. A group of emerging crisis care models with varying functions within service systems are increasingly prevalent: they have potential to offer greater choice and flexibility in managing crises, but an evidence base regarding impact on service user experiences and outcomes is yet to be established.

## Background

Developing and implementing alternatives to acute hospital admission has for several decades been a highly salient aim in mental health service development, endorsed by policy makers, service planners and service users (1). Inpatient care is costly, and potential harms include loss of rights and freedoms, stigma, institutionalisation and development of unhelpful coping strategies (2). Although inpatient care is sometimes the most appropriate option, most service users prefer services which are less institutional (3).

Three main alternatives to an acute inpatient admission have developed since the mid-20^th^ century, each giving rise to a substantial research literature (4). Crisis resolution teams (CRTs) are multidisciplinary teams that provide rapid assessment and intensive home treatment for a limited period during a crisis, and have become almost universal in English catchment areas since being mandated in the NHS Plan in 2000 (5). CRTs have also been widely implemented in Norway (6) and Germany (7). The second alternative, acute day units (ADUs) (traditionally known as acute day hospitals), provide a structured programme of activities and support on a group and individual basis for a time-limited period during a crisis. A review in 2011 found that for some service users, ADUs were a viable alternative to inpatient admission with similar rates of readmission (8). More recent studies carried out in England found that people supported by ADUs had a positive experience (9) and similar readmission rates to service users of CRTs (10). Although ADUs can be found across Europe (11), they are much less widely implemented than CRTs in England (12). The third alternative, crisis houses, are residential services which provide short-term intensive support in the community (13). Crisis houses may be cheaper than inpatient care and service user satisfaction tends to be high (13), with comparable rates of readmission despite shorter length of stay than inpatient wards (14). As is the case for ADUs, we are unaware of national policies to implement the crisis house model in any country.

Despite the provision of CRTs and sporadic availability of ADUs and crisis houses in England, there has been widespread dissatisfaction with crisis care. A national survey conducted by the Care Quality Commission in 2014 found that only 14% of respondents felt they received appropriate support in a mental health crisis (15). The share of mental health budgets dedicated to acute hospital care remains high, and the nationwide implementation of CRTs does not seem to have resulted in consistent reductions in admission rates as envisaged when they were originally mandated (16), in high quality community crisis care being available and readily accessed (17), or in a reduction in overall rates of detention or ethnic inequalities regarding who is detained (18).

Dissatisfaction with access to and quality of crisis care in England has been a driver for a national service improvement programme, with a Crisis Care Concordat introduced in 2014 (19). The aim of the Concordat was to facilitate local innovation, which was already occurring in some areas as a response to identified gaps in service provision (20). A national survey by our group in 2016 revealed wide variations in the provision of crisis services, and the emergence of several new models across the country (21). Grey literature (19, 22-25), including policy documents and voluntary sector reports, have documented some of these new models including “crisis cafés” (also often referred to as “safe havens” or “recovery cafés”): walk-in services allowing informal assessment; stand-alone community crisis assessment teams; and 24-hour crisis lines established to improve access and care provision.

Given the high priority attached to reductions in hospital admissions, especially involuntary, and improving mental health crisis care in England and internationally, there is considerable interest in these emerging models of crisis care (1). While descriptions and/or preliminary evaluations of some new models of mental health crisis care can be found in the literature, e.g. for police street triage teams (26) and psychiatric decision units (PDUs) (27), it is striking that several new models remain almost undocumented in the peer-reviewed research literature, and little is known about the extent and variation of their implementation in England. In general, they have developed according to local need, rather than via the process of theory building and iterative testing advocated in guidance such as the Medical Research Council (MRC) framework (28). If new models are to be recommended across England and beyond, clear definitions of their purposes and key components, is required as a prelude to acquiring evidence on their effectiveness and cost-effectiveness.

We conducted a national survey of mental health community crisis care services for all age groups in England in 2019, to identify and describe these emerging crisis service models. In this paper, we focus on the survey data regarding crisis care for adults of working age or all ages. The research questions were:

I. What are the characteristics of the new models of crisis care in England?
II. How widely disseminated are these new models of crisis care, and can trends over time be discerned when comparisons are made with a 2016 survey in which some preliminary information about these models was acquired?

## Methods

### Design and participants

From April - December 2019, we conducted a cross-sectional national survey of crisis care in England. The basic unit for our data collection was the catchment area of CRTs. Following government mandates, almost every area in England is served by such a team, with most mental health Trust catchment areas sub-divided by several CRTs (21). Our first choice of participant for each area was the CRT manager, as staff in this role are normally required to have a very good familiarity with crisis services in their local catchment areas. We asked participants to complete each question regarding local service provision on 31 March 2019. As a first step in carrying out the survey, we identified and mapped all CRTs in England, including CRTs specialising in children and young people, adults of working age or all ages, and older adults and people with dementia. We defined a CRT as a service which provides time-limited (usually up to around a month) acute care to people in their homes who are experiencing a mental health crisis, with the aim of averting hospital admission. We excluded generic services in which an acute response was one of several service functions, for example a general children and young people mental health service that also provides enhanced care to their service users when in crisis.

### Measures

Development of the survey was led by experts in crisis care research with relevant experience in the development of data collection tools (SJ and BLE; (21, 29, 30)) and was informed by iterative feedback from a working group set-up specifically for this study. This working group included people with relevant lived experience of using services or supporting those who do, senior practitioners and managers currently or recently working in CRTs, relevant policy officials, and other academic researchers with expertise in acute care from a range of disciplinary backgrounds. The survey was delivered via UCL Opinio, a tool for designing and hosting surveys online. The survey was designed so that it could be completed online by the participant or via telephone with a researcher. It was tested by the working group and then piloted with managers from three separate CRTs.

The survey included structured and free text questions regarding the CRT the participant managed, and other local community crisis care services available in the CRT’s catchment area. These included the types of service identified as emerging innovative models in our preliminary 2016 survey ((21)), as well as from the grey literature including policy documents, and the authors’ and working group’s personal knowledge. Box 1 shows the models included, and the operational definitions used to determine whether services met inclusion criteria for the survey.

#### Box 1

**Models of crisis care and their definition**

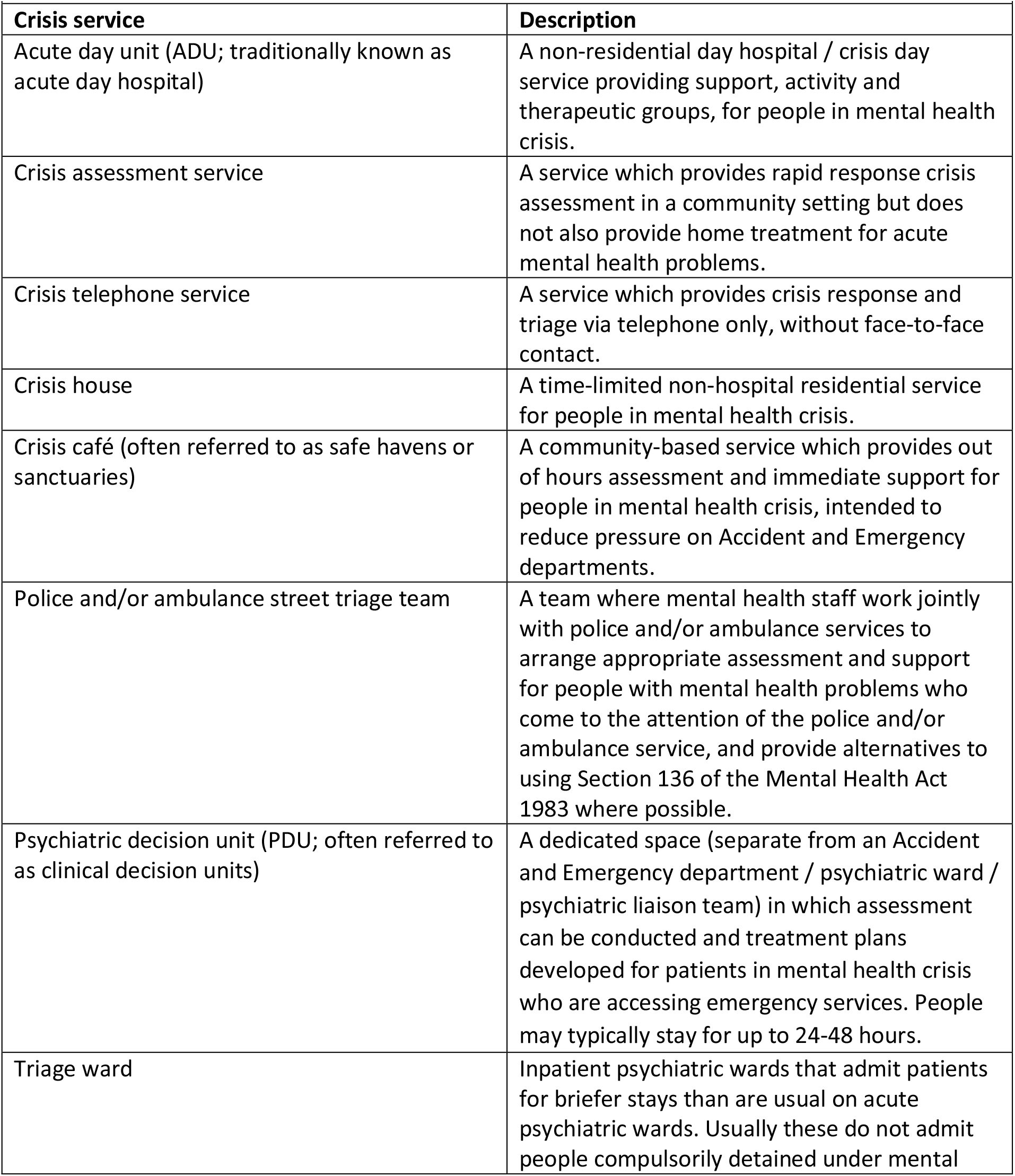
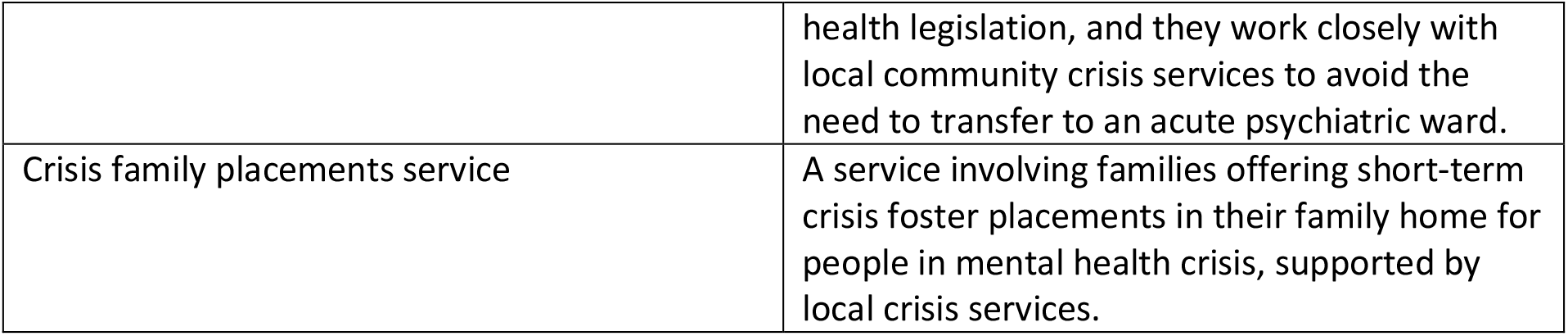

Participants were thus asked between 69 and 292 questions, depending on what services were provided locally. Each question included a ‘Not sure’ or ‘Don’t know’ response to avoid the participant guessing at answers. The survey took around 45 minutes to complete: a copy is included in the supplementary materials.

### Mapping

An initial list of CRTs in England serving all age groups was generated from four sources: i) a crisis care survey carried out by the research team in 2016 (21), ii) a separate survey carried out by NHS England in 2018-2019, iii) Freedom of Information requests sent to NHS Clinical Commissioning Groups (in the UK, under the Freedom of Information Act 2000 (31), members of the public have the right to request information held by public authorities); and iv) NHS Trust websites. Mapped CRT services were then contacted to confirm eligibility (i.e., that the primary remit of the service was to provide brief, intensive home treatment to people experiencing acute mental health problems, and the service was operational on 31 March 2019), contact details of the service manager or a senior member of staff which the survey invitation should be sent to, and to check details of other local CRTs.

Potential participants were sent an information sheet and a letter of support from the Department of Health and Social Care. The survey was completed by consenting participants online or via telephone interview with a researcher. Alternative respondents were sought where the person originally invited declined or did not respond.

### Data validation and cleaning

Where there was missing data, second respondents nominated by our initial respondent were contacted and asked to complete these questions. Researchers (CDL, BLE, LC and RS) then checked remaining gaps and inconsistencies in the data, with reference to available public information including Trust websites and a directory of voluntary sector crisis services provided from a recently completed study (20). All participants and NHS Trust Chief Executives were then contacted by the study team and invited to check and correct any remaining inaccuracies. Where information could not be confirmed or discrepancies resolved, data was treated as missing.

### Data analysis

This study reports descriptive results using Microsoft Excel and compares data from the current survey with a similar survey carried out in 2016 (21).

### Ethics

Our survey, commissioned by national policy-makers to understand current service provision, met national guidelines (32) for a service evaluation which did not require review from an ethics committee. We consulted Noclor, the research support service overseeing research governance for several NHS Trusts in North London to check this.

## Results

We mapped 200 CRTs serving working age adults or adults of all ages provided by 59 NHS Trusts and one independent health care provider. The survey opened 1 April 2019 and closed 18 December 2019. It used 31 March 2019 as the reference point for all questions. 184 of the 200 adult CRHTTs in England (92%), completed the survey. Completion was defined as having answered questions relating to the CRHTT and crisis assessment and telephone services. Only one NHS Mental Health Trust did not provide an adult CRT, compared to two in 2016 (21): this Trust is not included in this paper, which reports service provision at CRT catchment area-level. We also mapped 39 children and young people CRTs and 49 older adults/dementia CRTs.

Table 1 shows the types of crisis service available to the public for each adult CRT catchment area where there was a response to the survey, and compares this to data collected in the 2016 survey (21). A single crisis service may serve more than one adult CRT catchment area, and there may be more than one crisis service of each type in a local CRT catchment area. Therefore, this table does not show the total number of different crisis services in the country.

**Table 1:** Total mapped CRTs and the availability of other types of crisis service in 2016 and 2019.

|  |  |  |
| --- | --- | --- |
|  | **N (%) of adult CRTs with access to other crisis service models** |  |
| **Crisis service** | **2016 survey**<br>**(198 mapped adult CRTs)** | **2019 survey**<br>**(200 mapped adult CRTs)** |
| Children and young people CRT | 91/182 (50%) | 84/200 (42%) |
| Older adult/ dementia CRT | 77/183 (42%) | 73/200 (37%) |
| A separate crisis assessment service | 59/184 (32%) | 71/185 (38%) |
| A separate crisis telephone service | 132/184 (72%) | 115/184 (63%) |
| Psychiatric decision unit | Data not available | 30/182 (16%) |
| Triage ward | Data not available | 7/174 (4%) |
| Crisis house | 85/185 (46%) | 85/180 (47%) |
| Acute day unit | 40/185 (22%) | 20/182 (11%) |
| Crisis family placement scheme | Data not available | 5/174 (3%) |
| Crisis café | 28/185 (15%) | 52/180 (29%) |
| Police and/or ambulance street triage | 106/184 (58%) | 118/182 (65%) |

Table 1 illustrates that the level of provision of crisis houses in England has remained constant since 2016, with a large minority of CRTs having access, including gatekeeping access, to this form of crisis care. Access to acute day units has declined since 2016, while trends towards more widespread provision of other crisis care models can be observed: crisis assessment teams, crisis cafés and police and ambulance street triage teams. Other crisis models not included in the 2016 survey are also available in a minority of areas: Psychiatric Decision Units, inpatient triage wards and crisis family placements.

In the section below, we use our survey results to provide a structured summary of the core characteristics of each of these service models and variations in how they are organised. We also provide a case example of each model, using publicly available information. Further detail about the characteristics of each model is provided in the Supplementary Materials (Tables 1 to 8).

### Crisis assessment services

In 38% of CRT catchment areas, a separate crisis assessment team has been established, providing rapid assessment and referral on to other acute mental health services where appropriate. Whereas the original CRT model in England involves delivery of both rapid crisis assessment and intensive home treatment by the same team, in areas with crisis assessment services, these two functions are now provided by separate crisis assessment and crisis home treatment teams. Around two-thirds of crisis assessment services were operating 24-hours (48/70, 69%) and three-quarters accepted self-referrals from any member of the public as well as from professionals (53/71, 75%).

The First Response Service provided by Cambridge and Peterborough NHS Trust is an example of such a service (33). The service offers responsive, face-to-face crisis assessment in the community throughout their catchment area. It is open 24/7 and people can self-refer via NHS 111 option 2 (a national healthcare telephone triage service). They also accept referrals from anyone acting on behalf of the person in crisis, such as carers, GPs, and ambulance and police staff. The introduction of the First Response Service has been supported by the development of two crisis cafés, provided by a voluntary sector organisation and offering a range of support, including peer support.

### Crisis telephone services

A majority of CRTs had a separate dedicated crisis telephone service providing a response within their catchment area and triaging to other services in 2019 (115/184, 63%) but access to this type of crisis service had reduced since 2016 (132/184, 72%), possibly reflecting the increase of crisis assessment teams offering open access to face-to-face assessment. Most crisis telephone services were provided by the NHS (103/115, 91%), accessible by any member of the public seeking help within their catchment area (92/115, 80%) and available 24-hours (83/114, 73%).

The Single Point of Access service for Central and North West London NHS Trust (34) is available 24/7 via telephone and email. It can be accessed by the person in crisis or by someone on their behalf, including family, carers and other healthcare professionals such as GPs. Staff assess the urgency of the presented mental health problem and triage appropriately, which can include making appointments with other mental health services. Anyone who has concern for someone can also contact the service for advice and signposting.

### Crisis houses

Almost half of adult CRTs had access to a local crisis house (85/180, 47%), a 24 hour staffed community premises where people in crisis could go for a short stay. Just over half of CRTs who told us there was access to a local crisis house reported they had exclusive referral rights to the service so that all access was via this route (46/85, 54%). A minority reported the crisis house accepted self-referrals (14/85, 16%). Almost all CRTs in areas with crisis houses could ‘always’ or ‘usually’ access a crisis house bed when needed (74/83, 89%), and 43% told us they could arrange an admission within four hours (35/82).

Lowther Street Crisis House in Cumbria (35) is provided by the Richmond Fellowship, a national mental health charity. It is the only crisis house in Cumbria and accepts referrals 24/7 from other healthcare services. They have six places and a minimum of two staff on duty at all times, with a nurse available on call. The service aim is to provide an alternative to an acute hospital admission, in a homely, community setting.

The Drayton Park Women’s Crisis House, provided by Camden and Islington NHS Foundation Trust, is another example of a crisis house (36). They have 12 places available to women, including rooms where mothers can be admitted with their children. The service accepts self-referrals, and the staff, who are all women, are trained to work with women who may have experienced or are currently experiencing trauma such as sexual, physical or emotional abuse. The length of stay is initially seven days, but this can be extended up to a maximum of four weeks. Past and current residents are also invited to a weekly support group.

### Crisis cafés

The main remit of a crisis café is to provide a place other than hospital emergency departments where people in crisis can go to for support and signposting to other crisis services. They are often referred to as ‘safe havens’ or ‘recovery cafés’ and are typically provided by voluntary sector organisations. Such a service was available in around a third of adult CRT catchment areas in 2019 (52/180, 29%), up from 15% in 2016 (28/185). In most catchment areas, at least one of the crisis cafés was provided by the voluntary sector (40/48, 83%) and had a crisis café open seven days a week (32/48, 67%) for at least four hours, typically out-of-office hours (i.e. 5pm to 9am). Most allowed members of the public to attend without an appointment (32/47, 68%), and around a fifth were service user led (7/36, 19%).

The Bexley Crisis Café is provided by the mental health charity, Mind, and is located in south-east London (37). They are open 6-10pm seven days a week and adults over 18 can drop-in without an appointment to access mental health support and advice, including signposting to appropriate services. They aim to provide an environment which feels safe and welcoming for people in distress, and to help people feel less isolated.

Dial House in Leeds is provided by a partnership between Leeds Survivor-Led Crisis Service and Touchstone, offering a place of sanctuary in a homely environment (38). It is open 6pm-2am on Monday, Wednesday, Friday, Saturday, and Sunday, and people in crisis access the service by calling on the day that they wish to visit. They provide transport to and from the service and have a family room so parents can visit with their children.

### Acute day units (ADU)

Only 20 (11%) CRTs had access to an ADU, a day service where people in crisis could be admitted for a programme involving a mixture of therapies, activities and social contact. Managers of seven teams (35%) told us they had exclusive referral rights to the unit. Eight (40%) could ‘always’ access a place when needed and another 11 (55%) could usually access a place. Most could arrange a place within 24 hours (12/20, 60%).

The Fennell Acute Day Treatment Service in Coventry is provided by Coventry and Warwickshire Partnership NHS Trust (39). The service aims to provide an alternative to hospital admission and support early discharge from hospital, with a focus on crisis resolution and recovery. They provide individual and group focussed support including physical health assessments, signposting and vocational groups such as gardening and sewing.

### Police and/or ambulance street triage

Street triage services are available in about two thirds of CRT catchment areas (118/182, 65%). At least three different models of street triage have been implemented in England: i) two-fifths operated as a mobile unit with a mental health worker and a police officer or paramedic (44/109, 40%), attending calls where there was a suspected mental health element ; ii) around one-fifth had a mental health worker stationed at a call centre who could go out with a police officer or paramedic to calls when required (23/109, 21%); iii) just under a fifth told us that a mental health worker was stationed at the call centre and could provide phone advice only (19/109, 17%). The remainder selected ‘other’ (23/109, 21%). A quarter of CRTs who told us there was a local street triage team reported it was a 24-hour service (23/93, 25%).

The Teeside Street Triage, a partnership between Tees, Esk, and Wear Valley NHS Foundation Trust and Cleveland Police, is available daily from 4pm until midnight (40). When a police officer encounters someone they think may have a mental health need, they contact a team of mental health nurses who arrive at the location and carry out an assessment. The aim of the service was to reduce admissions under Section 136 of the Mental Health Act 1983, and provide people with mental health problems with more appropriate support. The team of nurses also provides training to Cleveland Police officers to increase mental health awareness.

### Psychiatric decision unit (PDU)

PDUs are dedicated spaces co-located with or accessed via a general hospital emergency department where a mental health assessment can be conducted and a support plan put in place for people who are in crisis. The majority of CRTs with a local PDU told us this unit was based in a psychiatric hospital (21/30, 70%), almost all were 24-hour (27/28, 96%), and around four-fifths provided reclining seats rather than beds (23/28, 82%).

The Sheffield Decision Unit, provided by Sheffield Health and Social Care NHS Foundation Trust, aims to offer a safe and comfortable place for people in crisis to be assessed (41). It is located near, but is separate from, the Northern General Hospital Accident and Emergency department. It is designed for short stays with a maximum stay of 48 hours. There are male and female lounge areas with recliners, no beds, and private assessment rooms.

### Triage wards

Triage wards are assessment wards with shorter stays than acute wards and work closely with community crisis services with the aim avoiding referral to an acute ward. They usually do not admit patients detained under the Mental Health Act 1983. In 2019, only seven out of 174 (4%) CRTs had access to a local triage ward. The median maximum length of stay was 72 hours and the median number of beds on the ward was 18.

The Swift Assessment for the Immediate Resolution of Emergencies (SAFIRE) Unit in Manchester, provided by Greater Manchester Mental Health NHS Foundation Trust, offers rapid assessments of people in mental health crisis with the aim of identifying the most appropriate form of support (42). Although this may entail transfer to an acute inpatient ward, the intention is to find an alternative to inpatient admission, which will usually mean a transfer to the CRT or to the community mental health team. The unit has nine beds and is available 24/7.

### Crisis family placement schemes

Crisis family placement schemes provide people in crisis with a trained and supported host family. Only five out of 174 CRTs (3%), all in the same NHS Trust, had access to a crisis family placement scheme for adults in 2019. This was an NHS-run service. The CRTs reported they could usually access a place when needed, and that placements could typically be arranged within 24 hours. A typical length of stay with the host family was just under four weeks.

The Hertfordshire Host Families Scheme, provided by Hertfordshire Partnership University NHS Foundation Trust, is intended to offer a welcoming family environment for a person in crisis as an alternative to hospital admission or to support an early discharge (43). Guests are encouraged to take part in daily life such as cooking meals and walking the dog. Host families and guests are supported by the local CRTs and the families receive a payment to cover the costs of having a guest. Guests usually stay for between three to six weeks.

### Crisis care systems

As Table 1 illustrates, there is wide variation in the local provision of different crisis care models, this leads to variation between local crisis care systems. Box 2 presents three examples of local crisis care systems, at the NHS Trust level. These do not represent defined types of system, but illustrate the extent and nature of variation within local crisis care systems.

#### Box 2

**Crisis care system examples**

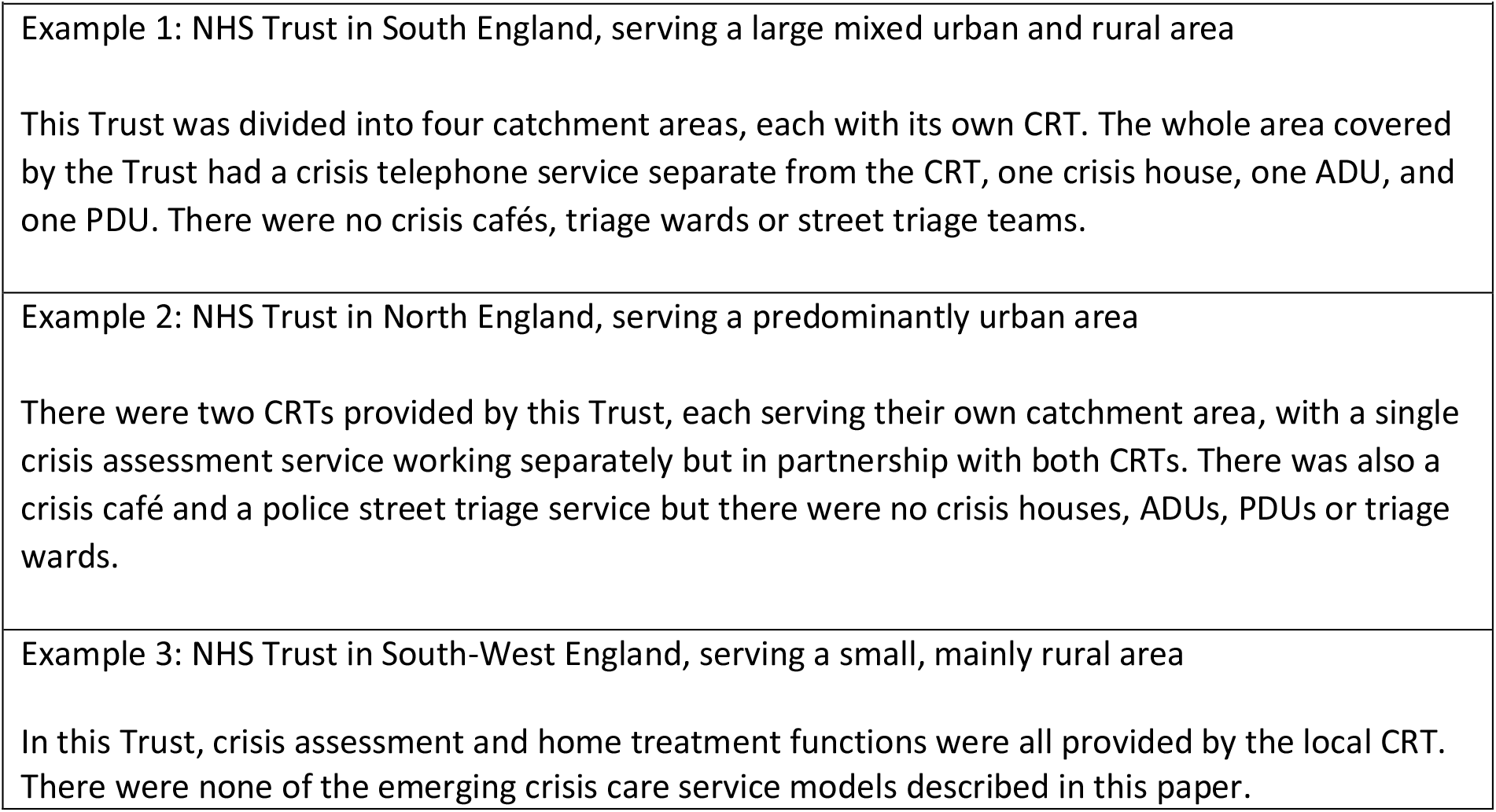
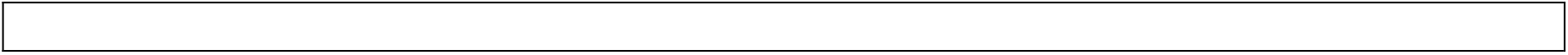

## Discussion

New models of crisis care have become widespread in England, but there is wide variation in local provision meaning some people have access to many alternative forms of community crisis care whilst others have access to just the crisis resolution teams that have been standard in the UK over the past 20 years, offering assessment, and where feasible, home treatment. The national implementation of CRTs following the NHS Plan (2000) (5) provided a local dedicated service whereby people acutely unwell could be assessed and treated in the community and in their own homes. However, gaps and shortfalls in quality have continued to be identified in the acute care system despite this national implementation, resulting in the past ten years in the proliferation of specialised crisis care models documented in our paper. This has potential to provide greater choice, flexibility and role clarity but may also come with risks of discontinuity in care, greater complexity and difficulty in navigating systems. Whole-system evaluation may be required to fully understand the impact of this change.

Furthermore, there does not appear to be a link between the extent of evidence and adoption of new models of crisis care. For example, crisis cafés have increased the most rapidly between 2016 and 2019 of all the emerging models but there is minimal evidence regarding the effectiveness of crisis cafés, whereas ADUs have a substantial body of evidence (8-10, 12) but have declined over the same period. This is inconsistent with aspirations that policy should be rooted in evidence in order to make best use of resources and maximise safety, acceptability and effectiveness of services. Thus, evidence now needs to be acquired regarding impacts of previously untested crisis care models’ safety, service users’ experiences and outcomes, and their impact on resource use in the system as a whole. Availability of a greater range of models and pathways in crisis care systems has potential to increase choice and the range of needs that can be successfully addressed, but we now need to understand which models work best for whom and under what circumstances, and whether any significant risks or unintended harms are involved.

For example, stand-alone crisis assessment services may be able to respond more quickly and flexibly to initial help-seeking than a traditional CRT which provides both assessment and home treatment, and are especially attractive in areas where there is a range of crisis care alternatives. However, a separate assessment team may take resource away from the home treatment team and reduce flexibility by allowing for less sharing of resource between crisis assessment and home treatment. It is also unclear how it affects the experience of the person in crisis. One of the criticisms levelled at the traditional CRT model is that the service user may be seen by several different healthcare clinicians during their brief time with the CRT, hampering the continuity of care and the formation of therapeutic relationships (44). This may be more likely if the person is assessed and treated by two different teams. Evaluation of this model is needed to better understand its impact on the system and service users. Crisis cafés which offer non-clinical forms of support in a less formal setting than a hospital emergency department may facilitate earlier help-seeking, for example for people who may be at risk of harming themselves if they become severely distressed. However, they may be less well equipped than hospital settings to provide swift access to physical tests or treatment or a full clinical assessment, with potential safety risks if widely used as a first point of crisis support. Psychiatric decision units may reduce pressure on emergency departments, but have also been criticised as holding distressed service users for extended periods in uncomfortable surroundings, for example with reclining seats rather than beds (45). Thus, it cannot be assumed that no unintended negative consequences result from new models.

## Limitations

This survey was carried out in 2019, and as the changes since 2016 indicate, this is an area of health care which changes quickly and so may not provide an accurate picture of current provision. The COVID-19 pandemic and its wide-reaching impact on mental health care is likely to have further accelerated changes in crisis care, with reports of new crisis assessment services being rapidly introduced (46, 47). This study does however illustrate how crisis care systems have continued to change since the implementation of CRTs. Our focus is only on England, so no conclusions can be drawn about whether new models described are also found internationally.

Primary respondents in this survey were usually the manager of the CRT, who despite their signposting and triage role may not have a fully comprehensive knowledge of local crisis care. Although wide-ranging, our survey may have failed to elicit information about unusual local crisis services which we did not ask about directly – for example, a chaplaincy team which patrols a local area well known for suicide attempts and tries to dissuade people from their planned course of action (48). We did however apply rigorous methods to ensure the accuracy of the data, including having ‘Not sure’ or ‘Don’t know’ response options to all survey questions to avoid participants guessing at answers, checking data with nominated second respondents where possible, and checking the data against a directory of voluntary care services (20).

Finally, the survey asked only about organisation and not about content of care in crisis care services. We provided brief operational definitions, and so there should be core features which are common within the different service models. However, there may still be considerable variation within these models. For example, a national survey of PDUs in England found wide variety in how these services are structured and what activities they provide (27).

### Research implications

Several directions for further research are suggested by our findings. First, evaluations are needed of emerging crisis care models, including new services implemented during the COVID-19 pandemic. These evaluations should focus on service user outcomes and experiences, and consider the potential of unintentional harm caused by the model. For example, crisis cafés provide great potential but it is currently unknown whether this usually non-professional setting is appropriate for managing mental health crises. Also, economic analyses are needed to ensure that models make effective and efficient use of limited resources. Second, we did not investigate the role of service users and carers in developing and providing leadership in crisis services, although there are examples where this is reported to work well (e.g. Dial House in Leeds, REF). Future research which helps better understand the role of people with lived experience in the commissioning, development, and delivery (as peer support workers), of emerging crisis care services will help inform future planning of services. Third, well-established crisis care models such as crisis houses and acute day services have been described and evaluated in a range of higher income countries. A task for further research is to establish whether the emerging models described in the current paper have also developed in other countries, how they fit into different crisis care systems if so, and what outcomes, service user experiences and risks may be associated with them in other contexts. Fourth, it is unclear whether there are any geographical or socio-economic factors associated with the current provision of innovative crisis care service models. It is important that if these new service types do provide favourable outcomes that certain areas are not left behind, potentially exacerbating social and health inequalities. Fifth, research should explore the possibility of developing a typology of crisis care systems, and if different definable systems do exist, they should also be evaluated. Crisis care systems are likely to be complex and vary greatly but a broader understanding of the wider system is needed to fully understand the role and impact of individual services and the contexts in which they perform best.

### Implications for policy and practice

Unlike the NHS Plan 2000 which provided a very prescriptive, top-down national overhaul of mental health crisis care, recent service developments have been locally driven and varied, usually without robust evaluation. This creates a challenge for national policymakers and local commissioners to know what best to promote and provide. However, we identify four implications for policy and practice from our study. First, there is no obvious justification for the apparent decline in the provision of evidence-based community crisis services such as ADUs. We suggest policy makers and service planners should prioritise providing models such as ADUs and crisis houses where there is substantial evidence of benefit, and also that models can be safely implemented. Second, development of new models of care may divert attention from improving quality and service user experiences in existing services, such as crisis resolution teams. Kindness and compassion, attributes that are both highly valued by people in crisis, are often lacking in mental health crisis care (15). Furthermore, in a recent study of 75 CRTs, none was found to be operating at high fidelity to the service model (17), but improvement initiatives can increase model fidelity and improve service outcomes (49). It is important that innovation in service models does not divert all resources and attention from efforts to maintain and improve the quality of care delivered within existing services. Third, our survey illustrates high levels of innovation and variation in local community crisis care systems. This is likely to have increased during the COVID-19 pandemic, which has caused widespread changes to mental health care (46). Given the lack of a robust evidence base regarding many of the emerging models described in our paper, and the high levels of service user dissatisfaction with crisis care, there is a clear case for any further restructuring of crisis care to be co-produced from the start in order to meet service users’ needs in a local context. Fourth, the variation in service provision may reflect a lack of prescriptive policy directives (beyond providing CRT (5)) and initiatives like the Crisis Care Concordat (19) which explicitly encourage local innovation. This may be appropriate given the widely recognised need to improve mental health crisis care and lack of evidence to support many emerging models, but it risks creating unwarranted variation and post code lotteries in what types of care are available in any given area. In the absence of sufficient evidence to guide policy clearly, national and local commissioners should diligently audit the impact of new services and changes to local crisis care systems, which will in turn increase the evidence base and inform subsequent decision-making.

## Conclusions

Crisis care in England has continued to evolve since the nationwide implementation of crisis resolution teams in 2000, with a trend toward specialisation seemingly borne out of local need and creating substantial regional variation. There are new and emerging crisis care models with great potential to improve access and service user satisfaction, but the evidence base for some of these models is lacking, and we do not yet know how introducing these new models affects the broader local crisis care system. Research evaluating these models and the adaptations made since the start of the COVID-19 pandemic is needed urgently to inform future crisis care provision.

## Lived experience commentary

### Chris Lynch and Karen Persaud

Crisis Care Services (CCS) for users and carers are often considered the most traumatising and dehumanising aspect of Mental Health Services so we are pleased to see this research into emerging new models. From personal experience what happens when someone is at crisis point and what response they receive can dramatically affect the rest of their lives. A kind, compassionate, caring, and effective response followed by the right support can transform lives for the better.

The research highlights the massive variety that is offered, much of which seems to be built around what has historically been available, what funding has been obtained and which direction commissioners have taken recently. It still very much feels like a system built up around things being done to people and for people rather than things done with people and the scope did not capture details of treatments provided (e.g., psycho-social elements and whether crisis planning played a role in any aspect). Also missing from the picture was the role of carers despite being a critical element of the cycle. These aspects need further exploration for impact and efficacy.

Co-production was touched on but no mention of whether patients/service users and carers had a role in the development. Given the ambition is creating a more inclusive and responsive service, service users and carers are a critical element of the development of CCS and this involvement must be measured as part of the whole picture. It is concerning (but not surprising) that evidence suggests CRTs sometimes do not work effectively to reduce admission rates. We need to know why. Customer feedback and satisfaction surveys would be a useful element of future data gathering exercises.

We want to see more around the quality of services and outcomes, ideally co-evaluated by people that have used them or cared for people that have. We think it’s important that the research is co-produced in the future and services co-delivered, otherwise, we’ll keep getting what we’ve always got.

We support the paper’s recommendation for a new body of research to delve further into these emerging models and would advocate for inclusion of services outside the NHS and for any future work to seek to better understand what CCS means for marginalised groups (e.g., Afro-Caribbean males).

## Data Availability

The dataset used and analysed during the current study are available from the corresponding author on reasonable request.

## Abbreviations

ADU: Acute Day Unit
CCS: Crisis Care Services
CRT: Crisis Resolution Team
PDU: Psychiatric Decision Unit

## Declarations

### Ethics approval and consent to participate

The study was reviewed and approved as a service evaluation by Noclor, the research support service overseeing research governance for several NHS Trusts in North London. As a service evaluation rather than research in which no personal data were collected, the survey did not require review and approval from a research ethics committee. Wherever respondents directed us to local R&D or audit committees, we followed Trusts’ local governance procedures.

### Consent for publication

Our plans to publish findings from this study was explained to participants in the Introduction to the survey. This text can be viewed in the copy of the survey included in the supplementary materials.

### Authors’ contributions

SJ and BLE made the initial plan for the study. All authors contributed to protocol development. SJ drafted the survey, all authors commented, and CDL developed the online version for UCL Opinio. CDL and BLE led on mapping services and data collection. CDL, JHS, NL, LSR and RS mapped services, invited managers to complete the survey, and conducting telephone interviews with participants. Data cleaning was led by CDL and performed by CDL, BLE and LC. CDL prepared and finalised the results tables. CDL, BLE and SJ drafted, and all authors contributed to and approved the final manuscript.

## Acknowledgements

The authors would like to thank the participants for responding to our invitations and taking the time to complete the survey. We are grateful to mental health policy colleagues in the Department of Health and Social Care and NHS England for their help in planning this survey and with mapping and approaching CRTs to take part.

## References

1. Lloyd-Evans B, Johnson S. Community alternatives to inpatient admissions in psychiatry. World Psychiatry. 2019;18(1):31–2.

2. Bowers L, Chaplin R, Quirk A, Lelliott P. A conceptual model of the aims and functions of acute inpatient psychiatry. J Ment Health. 2009;18(4):316–25.

3. Farrelly S, Brown G, Rose D, Doherty E, Henderson RC, Birchwood M, et al. What service users with psychotic disorders want in a mental health crisis or relapse: thematic analysis of joint crisis plans. Soc Psych Psych Epid. 2014;49(10):1609–17.

4. Johnson S, Totman J, Hobbs L. Crisis and emergency services. In: Thornicroft G, Szmuckler G, Mueser K, Drake R, editors. Oxford Textbook of Community Mental Health. Oxford: Oxford University Press; 2011. p. 118–28.

5. Department of Health. The NHS Plan: a plan for investment, a plan for reform. 2000 1 July 2000.

6. Johnson S. Crisis resolution and home treatment teams: an evolving model. Advances in Psychiatric treatment. 2013;19(2):115–23.

7. Lambert M, Karow A, Gallinat J, Ludecke D, Kraft V, Rohenkohl A, et al. Study protocol for a randomised controlled trial evaluating an evidence-based, stepped and coordinated care service model for mental disorders (RECOVER). BMJ Open. 2020;10(5):e036021.

8. Marshall M, Crowther R, Sledge WH, Rathbone J, Soares-Weiser K. Day hospital versus admission for acute psychiatric disorders. Cochrane Database Syst Rev. 2011(12):CD004026.

9. Morant N, Davidson M, Wackett J, Lamb D, Pinfold V, Smith D, et al. Acute day units for mental health crises: a qualitative study of service user and staff views and experiences. Bmc Psychiatry. 2021;21(1).

10. Lamb D, Steare T, Marston L, Canaway A, Johnson S, Kirkbride JB, et al. A comparison of clinical outcomes, service satisfaction and well-being in people using acute day units and crisis resolution teams: cohort study in England. Bjpsych Open. 2021;7(2).

11. Kallert TW, Glockner M, Priebe S, Briscoe J, Rymaszewska J, Adamowski T, et al. A comparison of psychiatric day hospitals in five European countries: implications of their diversity for day hospital research. Soc Psychiatry Psychiatr Epidemiol. 2004;39(10):777–88.

12. Lamb D, Davidson M, Lloyd-Evans B, Johnson S, Heinkel S, Steare T, et al. Adult mental health provision in England: a national survey of acute day units. BMC Health Serv Res. 2019;19(1):866.

13. Lloyd-Evans B, Slade M, Jagielska D, Johnson S. Residential alternatives to acute psychiatric hospital admission: systematic review. Br J Psychiatry. 2009;195(2):109–17.

14. Byford S, Sharac J, Lloyd-Evans B, Gilburt H, Osborn DP, Leese M, et al. Alternatives to standard acute in-patient care in England: readmissions, service use and cost after discharge. Br J Psychiatry Suppl. 2010;53:s20–5.

15. Care Quality Commission. Right here, right now – help, care and support during a mental health crisis. 2015.

16. Jacobs R, Barrenho E. Impact of crisis resolution and home treatment teams on psychiatric admissions in England. Br J Psychiatry. 2011;199(1):71–6.

17. Lamb D, Lloyd-Evans B, Fullarton K, Kelly K, Goater N, Mason O, et al. Crisis resolution and home treatment in the UK: A survey of model fidelity using a novel review methodology. Int J Ment Health Nurs. 2020;29(2):187–201.

18. Paton F, Wright K, Ayre N, Dare C, Johnson S, Lloyd-Evans B, et al. Improving outcomes for people in mental health crisis: a rapid synthesis of the evidence for available models of care. Health Technol Assess. 2016;20(3):1–162.

19. HM Government. Mental Health Crisis Care Concordat Improving outcomes for people experiencing mental health crisis. 2014.

20. Newbigging K, Rees J, Ince R, Mohan J, Joseph D, Ashman M, et al. The contribution of the voluntary sector to mental health crisis care: a mixed-methods study. Health Services and Delivery Research. 2020;8(29).

21. Lloyd-Evans B, Lamb D, Barnby J, Eskinazi M, Turner A, Johnson S. Mental health crisis resolution teams and crisis care systems in England: a national survey. BJPsych Bull. 2018;42(4):146–51.

22. Mind. Listening to experience: An independent inquiry into acute and crisis mental health care. 2011.

23. Wood C. Listen and you might Learn. Pure Potential Scotland; 2016.

24. Wood C. Proposal for an Out of Hours Crisis Café for North East Glasgow. Pure Potential Scotland; 2017.

25. Surrey and Borders Partnership Foundation NHS Trust. ‘The Safe Haven’ Aldershot: Evaluation Report. 2014.

26. Reveruzzi B, Pilling S. Street Triage: Report on the evaluation of nine pilot schemes in England. UCL; 2016.

27. Goldsmith LP, Anderson K, Clarke G, Crowe C, Jarman H, Johnson S, et al. The psychiatric decision unit as an emerging model in mental health crisis care: a national survey in England. Int J Ment Health Nurs. 2021.

28. Medical Research Council. Developing and evaluating complex interventions: New guidance. 2019.

29. Johnson S, Gilburt H, Lloyd-Evans B, Osborn DP, Boardman J, Leese M, et al. In-patient and residential alternatives to standard acute psychiatric wards in England. Br J Psychiatry. 2009;194(5):456–63.

30. Lloyd-Evans B, Paterson B, Onyett S, Brown E, Istead H, Gray R, et al. National implementation of a mental health service model: A survey of Crisis Resolution Teams in England. International Journal of Mental Health Nursing. 2018;27(1):214–26.

31. UK Parliament. Freedom of Information Act 2000. 2000.

32. Medical Research Council (MRC) Regulatory Support Centre. Is my study research? 2017 [cited 2021 18/05/2021]. Available from: http://www.hra-decisiontools.org.uk/research/.

33. Cambridge and Peterborough NHS Foundation Trust. First Response Service (FRS) 2021 [Available from: https://www.cpft.nhs.uk/service-detail/service/first-response-service-frs-21/.

34. Central and North West London NHS Trust. Single Point of Access service 2021 [cited 2021 10/05/2021]. Available from: https://www.cnwl.nhs.uk/services/mental-health-services/adult-and-older-adult/single-point-access.

35. Richmond Fellowship. Lowther Street Crisis House in Cumbria 2021 [cited 2021 10/05/2021]. Available from: https://www.recoveryfocus.org.uk/find-services/crisis-support-service/lowther-street-crisis/?view=map-view.

36. Camden and Islington NHS Foundation Trust. Drayton Park Women’s Crisis House 2021 [cited 2021 10/05/2021]. Available from: https://www.candi.nhs.uk/services/drayton-park-womens-crisis-house-and-resource-centre.

37. Mind. Bexley Crisis Café 2021 [cited 2021 10/05/2021]. Available from: https://mindinbexley.org.uk/crisis-cafe.

38. Leeds Survivor-Led Crisis Service. Dial House 2021 [cited 2021 18/05/2021]. Available from: https://www.lslcs.org.uk/about-us/.

39. Coventry and Warwickshire Partnership NHS Trust. Fennell Acute Day Treatment Service in Coventry 2021 [cited 2021 10/05/2021]. Available from: https://www.covwarkpt.nhs.uk/service-detail/health-service/fennell-acute-day-treatment-service-756/.

40. Tees, Esk, and Wear Valley NHS Foundation Trust. Teeside Street Triage 2021 [cited 2021 10/05/2021]. Available from: https://www.tewv.nhs.uk/services/street-triage/.

41. Sheffield Health and Social Care NHS Foundation Trust. Sheffield Decision Unit 2021 [cited 2021 10/05/2021]. Available from: https://www.shsc.nhs.uk/services/decisions-unit

42. Greater Manchester Mental Health NHS Foundation Trust. Swift Assessment for the Immediate Resolution of Emergencies (SAFIRE) Unit in Manchester 2021 [cited 2021 10/05/2021]. Available from: https://www.gmmh.nhs.uk/safire-unit.

43. Hertfordshire Partnership University NHS Foundation Trust. Hertfordshire Host Families Scheme 2021 [cited 2021 10/05/2021]. Available from: https://www.hpft.nhs.uk/services/acute-and-rehabilitation-services/alternative-services-to-an-inpatient-stay/host-families-scheme/.

44. Morant N, Lloyd-Evans B, Lamb D, Fullarton K, Brown E, Paterson B, et al. Crisis resolution and home treatment: stakeholders’ views on critical ingredients and implementation in England. Bmc Psychiatry. 2017;17(1):254.

45. Hussain D. Mental health patient slept in a CHAIR for a week while waiting for a bed. Birmingham Live. 2017.

46. Johnson S, Dalton-Locke C, Vera San Juan N, Foye U, Oram S, Papamichail A, et al. Impact on mental health care and on mental health service users of the COVID-19 pandemic: a mixed methods survey of UK mental health care staff. Soc Psychiatry Psychiatr Epidemiol. 2021;56(1):25–37.

47. Royal College of Psychiatrists. Alternatives to emergency departments for mental health assessments during the COVID-19 pandemic. 2020.

48. National Suicide Prevention Alliance. Beachy Head Chaplaincy Team 2021 [cited 2021 18/05/2021]. Available from: https://nspa.org.uk/member/beachy-head-chaplaincy-team/.

49. Lloyd-Evans B, Osborn D, Marston L, Lamb D, Ambler G, Hunter R, et al. The CORE service improvement programme for mental health crisis resolution teams: results from a cluster-randomised trial. Br J Psychiatry. 2020;216(6):314–22.

