## Supplementary Materials 1 for "Emerging models and trends in mental health crisis care in England: a national investigation of crisis care systems"

### The NIHR Mental Health Policy Research Unit 2019 Survey of Crisis Care Systems in England

Supplementary Materials



Mental Health
Policy Research Unit
mhpru

### Table 1: Crisis assessment services

| | ***Yes*** | | ***No*** | | ***Unsure/ Missing*** |
|---|---|---|---|---|---|
| | ***n*** | ***%*** | ***n*** | ***%*** | ***n*** |
| ***Is there a separate crisis assessment service in your area?*** | 71 | 38 | 114 | 62 | 15* |
| ***24hr*** | 48 | 69 | 22 | 31 | 1** |
| ***Any member of public can self-refer*** | 53 | 75 | 18 | 25 | 0** |
| ***Anyone known to the service can self-refer*** | 59 | 83 | 12 | 17 | 0** |
| ***Can decide who should be treated by CRHTT without re-assessment*** | 53 | 76 | 17 | 24 | 1** |
| ***Is there any shared staffing between the crisis assessment service and any other local crisis service?*** | 35 | 50 | 35 | 50 | 1** |
| ***Is there any joint management between the crisis assessment service and any other local crisis service?*** | 47 | 68 | 22 | 32 | 2** |

*Number 'Unsure/Missing' out of all CRHTTs mapped (n=200). **Number 'Unsure/Missing' out of all areas with a crisis assessment service (n=71).

### Table 2: Crisis telephone services

| | *Yes* | | *No* | | *Unsure/ Missing* |
|---|---|---|---|---|---|
| | *n* | *%* | *n* | *%* | *n* |
| ***Is there a separate crisis telephone service in your area?*** | 115 | 63 | 69 | 38 | 16* |
| ***Provider*** | - | - | - | - | 2** |
| ***NHS*** | 103 | 91 | - | - | - |
| ***Voluntary*** | 7 | 6 | - | - | - |
| ***Local authority*** | 3 | 3 | - | - | - |
| ***Managed as part of another service*** | 69 | 62 | 42 | 38 | 4** |
| ***24hr*** | 83 | 73 | 31 | 27 | 1** |
| ***Any member of public can access*** | 92 | 80 | 23 | 20 | 0** |
| ***Can decide who should be treated by CRHTT without re-assessment*** | 30 | 26 | 85 | 74 | 0** |
| ***Can admit to hospital without reference to CRHTT*** | 9 | 8 | 106 | 92 | 0** |
| ***Is there any shared staffing between the crisis telephone service and any other local crisis service?*** | 34 | 32 | 73 | 68 | 8** |
| ***Is there any joint management between the crisis telephone service and any other local crisis service?*** | 50 | 46 | 58 | 54 | 7** |

*Number 'Unsure/Missing' out of all CRHTTs mapped (n=200). **Number 'Unsure/Missing' out of all areas with a crisis telephone service (n=115).

### Table 3: Crisis houses

| | Yes | | No | | Unsure/ Missing |
|---|---|---|---|---|---|
| | *n* | *%* | *n* | *%* | *n* |
| ***Is there at least one crisis house in your area?*** | 85 | 47 | 95 | 53 | 20* |
| ***Is there > one crisis house in your area?*** | 21 | 12 | 156 | 88 | 23* |
| ***The CRHTT has exclusive referral rights to a crisis house*** | 46 | 26 | 134 | 75 | 20* |
| ***People in crisis can self-refer to a crisis house*** | 14 | 8 | 166 | 93 | 20* |
| ***The CRHTT/ crisis assessment service can access a crisis house bed when needed?*** | - | - | - | - | 2** |
| ***Always*** | 25 | 30 | - | - | - |
| ***Usually*** | 49 | 59 | - | - | - |
| ***Sometimes*** | 7 | 8 | - | - | - |
| ***Rarely*** | 2 | 2 | - | - | - |
| ***How quickly can admission to a crisis house be arranged?*** | - | - | - | - | 3** |
| ***< 4 hours*** | 35 | 43 | - | - | - |
| ***< 24 hours*** | 43 | 52 | - | - | - |
| ***< 72 hours*** | 4 | 5 | - | - | - |
| ***Is there a local crisis house which is service user led?*** | 6 | 8 | 73 | 92 | 6** |
| **-** | ***n*** | ***Median*** | ***IQR*** | **-** | ***Unsure/ Missing*** |
| ***Minimum age?**** | 80 | 18 | 0 | - | 5** |
| ***What is the typical length of stay, in days?**** | 70 | 7 | 5 | - | 15** |
| ***What is the maximum length of stay, in days?**** | 80 | 7 | 7 | - | 5** |

*Number 'Unsure/Missing' out of all CRHTTs mapped (n=200). **Number 'Unsure/Missing' out of all areas with a crisis house (n=85). ***Calculated from the data regarding the first crisis house the participant told us about.

#### Table 4: Crisis cafés

| | Yes | | No | | Unsure/ Missing |
|---|---|---|---|---|---|
| | *n* | *%* | *n* | *%* | *n* |
| ***Is there at least one crisis café in your area?*** | 52 | 29 | 128 | 71 | 20* |
| ***Is there > one crisis café in your area?*** | 12 | 7 | 159 | 93 | 29* |
| ***Is at least one crisis café provided by:*** | - | - | - | - | 4** |
| ***Local authority*** | 2 | 4 | - | - | - |
| ***NHS*** | 6 | 13 | - | - | - |
| ***Voluntary sector*** | 40 | 83 | - | - | - |
| ***There is a crisis café open****** | - | - | - | - | **4 |
| ***Out of hours, Mon-Sun*** | 32 | 67 | - | - | - |
| ***Out of hours, Weekends*** | 9 | 19 | - | - | - |
| ***Out of hours, Weekdays*** | 1 | 2 | - | - | - |
| ***Office hours, Mon-Sun*** | 2 | 4 | - | - | - |
| ***Office hours, Weekends*** | 0 | 0 | - | - | - |
| ***Office hours, Weekdays*** | 1 | 2 | - | - | - |
| ***Other*** | 3 | 6 | - | - | - |
| ***Referral to crisis café*** | - | - | - | - | 5** |
| ***Can drop in without appointment*** | 32 | 68 | - | - | - |
| ***Can self-referral but need to book an appointment*** | 10 | 21 | - | - | - |
| ***Referral needed from other health service*** | 5 | 11 | - | - | - |
| ***Is there a local crisis café which is service user led?*** | 7 | 19 | 29 | 81 | 16** |

*Number 'Unsure/Missing' out of all CRHTTs mapped (n=200). **Number 'Unsure/Missing' out of all areas with a crisis café (n=52). ***'Out of hours' = at least open 4 hours between 5pm and 9am; 'Office hours' = at least open 4 hours between 9am and 5pm; 'Weekends' = at least 2 evenings between Thu-Sun; 'Weekdays' = at least 4 days between Mon-Fri.

### Table 5: Acute day units (ADU)

| | Yes | | No | | Unsure/ Missing |
|---|---|---|---|---|---|
| | n | % | n | % | n |
| ***Is there at least one ADU in your area?*** | 20 | 11 | 162 | 89 | 18* |
| ***Is there > one ADU in your area?*** | 3 | 2 | 177 | 98 | 20* |
| ***The CRHTT has exclusive referral rights to an ADU*** | 7 | 4 | 175 | 96 | 18* |
| ***People in crisis can self-refer to an ADU*** | 1 | 1 | 181 | 99 | 18* |
| ***The CRHTT/ crisis assessment service can access an ADU place when needed?*** | - | - | - | - | 0** |
| ***Always*** | 8 | 40 | - | - | - |
| ***Usually*** | 11 | 55 | - | - | - |
| ***Sometimes*** | 1 | 5 | - | - | - |
| ***Rarely*** | 0 | 0 | - | - | - |
| ***How quickly can admission to an ADU be arranged?*** | - | - | - | - | 0** |
| ***< 4 hours*** | 1 | 5 | - | - | - |
| ***< 24 hours*** | 11 | 55 | - | - | - |
| ***< 72 hours*** | 1 | 5 | - | - | - |
| ***< 1 week*** | 5 | 25 | - | - | - |
| ***> 1 week*** | 2 | 10 | - | - | - |
| ***Is there a local ADU which is service user led?*** | 1 | 5 | 18 | 95 | 1** |
| ***Is there any shared staffing between the ADU and any other local crisis service?*** | 2 | 10 | 18 | 90 | 0** |
| ***Is there any joint management between the ADU and any other local crisis service?*** | 6 | 32 | 13 | 68 | 1** |
| - | ***n*** | ***Med-ian*** | ***IQR*** | - | ***Unsure/ Missing*** |
| ***What is the typical length of time a patient can use this service, in days?*** | 17 | 14 | 16 | - | 3** |
| ***What is the maximum length of time a patient can use this service, in days?*** | 18 | 28 | 28 | - | 2** |

*Number 'Unsure/Missing' out of all CRHTTs mapped (n=200). **Number 'Unsure/Missing' out of all areas with an ADU (n=20). ***Calculated from the data regarding the first ADU the participant told us about.

#### Table 6: Police and/or ambulance street triage

| | Yes | | No | | Unsure/ Missing |
|---|---|---|---|---|---|
| | n | % | n | % | n |
| ***Is there a street triage in your area?*** | 117 | 65 | 64 | 35 | 19* |
| ***Is it 24hr?*** | 23 | 25 | 70 | 75 | 24** |
| ***What is the model of care*** | - | - | - | - | 8** |
| ***Mobile unit with mental health worker and police staff*** | 44 | 40 | - | - | - |
| ***Mental health worker based in service call centre – can attend calls*** | 23 | 21 | - | - | - |
| ***Mental health worker based in service call centre – phone advice only*** | 19 | 17 | - | - | - |
| ***Other*** | 23 | 21 | - | - | - |
| ***Is there any shared staffing between the police street triage and any other local crisis service?*** | 46 | 46 | 64 | 64 | 7** |
| ***Is there any joint management between the police street triage and any other local crisis service?*** | 61 | 55 | 49 | 45 | 7** |

*Number 'Unsure/Missing' out of all CRHTTs mapped (n=200). **Number 'Unsure/Missing' out of all areas with a police street triage (n=116).

### Table 7: Psychiatric decision units (PDU)

A PDU is a dedicated area (separate to an A&E department / psychiatric ward / psychiatric liaison team) in which assessment can be conducted and treatment plans developed for patients in mental health crisis who are accessing emergency services. People may typically stay for up to 24-48 hours in a PDU, which may be called a "Clinical Decision Unit" or other in some areas.

| | Yes | | No | | *Unsure/ Missing* |
|---|---|---|---|---|---|
| | *n* | *%* | *n* | *%* | *n* |
| ***Is there a PDU in your area?*** | 30 | 16 | 152 | 84 | 18* |
| ***Where is the PDU located*** | - | - | - | - | 0** |
| ***Psychiatric hospital*** | 21 | 70 | - | - | - |
| ***Acute hospital*** | 6 | 20 | - | - | - |
| ***Other*** | 3 | 10 | - | - | - |
| ***Is the PDU co-located with any other service?*** | 14 | 50 | 14 | 50 | 2** |
| ***Is it 24hr?*** | 27 | 96 | 1 | 4 | 2** |
| ***What is the accommodation on the PDU like?*** | - | - | - | - | 2** |
| ***Single room/ Partitioned area*** | 3 | 11 | - | - | - |
| ***Beds*** | 1 | 4 | - | - | - |
| ***Recliners*** | 23 | 82 | - | - | - |
| ***No overnight accommodation*** | 0 | 0 | - | - | - |
| ***Other*** | 1 | 4 | - | - | - |
| ***The CRHTT or crisis assessment service can refer*** | 20 | 69 | 9 | 31 | 1** |
| ***The psychiatric liaison service can refer*** | 26 | 90 | 3 | 10 | 1** |
| ***When can the PDU admit to hospital?*** | - | - | - | - | 1** |
| ***Without CRHTT or crisis assessment service input*** | 12 | 44 | - | - | - |
| ***Following phone discussion with CRHTT / crisis assessment service*** | 8 | 30 | - | - | - |
| ***Only following in person assessment of the patient by CRHTT / crisis assessment service*** | 7 | 26 | - | - | - |
| ***Other*** | 2 | - | - | - | - |
| ***Is there any shared staffing between the PDU and any other local crisis service?*** | 21 | 75 | 7 | 25 | 2** |
| ***Is there any joint management between the PDU and any other local crisis service?*** | 19 | 70 | 8 | 30 | 3** |
| *-* | *n* | *Median* | *IQR* | *-* | *Unsure/ Missing* |
| ***How long is the maximum length of stay, in hours?*** | 25 | 23 | 25 | - | 5** |
| ***What is the capacity of the PDU?*** | 23 | 6 | 1.5 | - | 7** |

*Number 'Unsure/Missing' out of all CRHTTs mapped (n=200). **Number 'Unsure/Missing' out of all areas with a PDU (n=30).



Mental Health Policy Research Unit mhpru

#### Table 8: Triage wards

An inpatient psychiatric ward which only accepts admissions for a time-limited period (not more than one week); typically does not admit people compulsorily detained under a MHA section; and works closely with local community crisis services to avoid the need to transfer to an acute psychiatric ward.

| | Yes | | No | | *Unsure/ Missing* |
|---|---|---|---|---|---|
| | *n* | *%* | *n* | *%* | *n* |
| ***Is there a triage ward in your area?*** | 7 | 4 | 167 | 96 | 26* |
| ***The CRHTT or crisis assessment service can refer*** | 6 | 86 | 1 | 14 | 0** |
| ***The psychiatric liaison service can refer*** | 5 | 71 | 2 | 29 | 0** |
| ***When can the triage ward admit to hospital?*** | - | - | - | - | 0** |
| ***Without CRHTT or crisis assessment service input*** | 3 | 75 | - | - | - |
| ***Following phone discussion with CRHTT / crisis assessment service*** | 1 | 25 | - | - | - |
| ***Only following in person assessment of the patient by CRHTT / crisis assessment service*** | 0 | 0 | - | - | - |
| ***Other*** | 3 | - | - | - | - |
| ***Is there any shared staffing between the triage ward and any other local crisis service?*** | 1 | 17 | 5 | 83 | 1** |
| ***Is there any joint management between the triage ward and any other local crisis service?*** | 0 | 0 | 6 | 100 | 1** |
| - | *n* | *Median* | *IQR* | - | *Unsure/ Missing* |
| ***How long is the maximum length of stay, in hours?*** | 5 | 72 | 0 | - | 2** |
| ***How many beds does it have?*** | 5 | 18 | 2 | - | 2** |

*Number 'Unsure/Missing' out of all CRHTTs mapped (n=200). **Number 'Unsure/Missing' out of all areas with a triage ward (n=7).

Mental Health Policy Research Unit
mhpru
