## Supplementary Materials 2 for "Emerging models and trends in mental health crisis care in England: a national investigation of crisis care systems"

### FINAL: NIHR MHPRU - Crisis care survey_28/03/2019

**Introduction**

By completing this questionnaire you will be helping with a national survey commissioned by the Department of Health and Social Care (DHSC).

The NHS Long Term Plan makes a commitment to increase investment in crisis pathways and there is strong support nationally and in many local services for integrated crisis pathways with a range of options that meet different needs and preferences. However, the evidence base regarding effective crisis systems limited. This survey will support national and local investment decisions over the next 5 years and inform an ongoing programme of research commissioned by the DHSC. This survey aims to complement the annual NHS England survey of mental health crisis teams, and has been developed with input from NHS England.

You have been asked to take part in this survey because you are the manager or a senior clinician working in one or more NHS mental health crisis services, which provide crisis assessment and/or intensive home treatment. If there is no stand-alone crisis assessment team or crisis home treatment team or equivalent in the Trust where you work, you have been asked to complete this survey as the manager of another NHS crisis service. We will ask you about availability of crisis services in your team and local area. The survey should take about 30 minutes to complete. If necessary, we will contact more than one respondent in each NHS Trust to obtain all the information we want about local crisis services. Please answer all the questions you can.

The survey is being carried out by a team of researchers based at University College London from the NIHR Mental Health Policy Research Unit. A summary of the findings will be written up in a report for the DHSC. In this report, and in any other publications resulting from this survey, you and the service you work in will be anonymised. We will provide the data from the survey to the DHSC and NHS England at the end of the survey. We will remove respondents' names from this data set so you will not be personally identified, but the team in which you work and other local crisis services will be identifiable.

We are aware that the names and functions of mental health crisis teams vary across England. We therefore recommend completing the survey as a telephone interview with a researcher, so any uncertainties over terms or definitions can be clarified immediately. However, you can complete the survey here online if you prefer and contact a researcher during the survey should you encounter any issues. Researcher contact details are provided at the bottom of each page.

If we do not hear anything from you, a member of the research team will get in touch to arrange a time when they can go through the questionnaire over the phone.

By pressing start, you confirm that you have read this information. You have also been sent a more detailed information sheet about the project when you were invited to take part – please contact the study researchers if you need another copy of this.

*Please complete the survey with how things were as of 31st March 2019, even if there have been changes since this date.*

**Instructions to complete the survey online**

After you have answered each question, press the 'next' button at the bottom of the screen to move to the next page or the 'back' button to return to any previous answers. You can also save

the survey and return to it at a later time by pressing ‘save’ then entering your email address and clicking on the link in the email you receive when you are ready to resume the survey.
Please note, if you have partially answered some questions but not the rest, the research team will be in touch by phone so that we can complete the survey.
If you prefer to complete the survey by phone, please contact one of our researchers. You can find their contact details at the bottom of this page. Please do not hesitate to contact us if you have any queries.
By pressing start, you confirm that you have read this information. You have also been sent a more detailed information sheet about the project when you were invited to take part – please contact the study researchers if you need another copy of this.
Thank you for your participation!

#### Respondent's information

**Q1: Respondent's name:**

**Q2: Respondent's job title:**

**Q3: Do you work in or manage more than one service?**

○ Yes ○ No

#### Respondent's information (cont.)

*Note: if you have answered/chosen item [1] in question 3, skip the following question*

**Q4: What is the name of the service you work in or manage?**

*Note: if you have answered/chosen item [2] in question 3, skip the following question*

**Q5: What are the names of the services you work in or manage?**
**Please complete the remainder of the survey in relation to the first service listed. We may later ask you about the other services you work in or manage.**

#### Respondent's information (cont.)

**Q6: Which NHS Trust provides your service?**

**Q7: What is the primary commissioning service or services (e.g. CCG) for this service?**

○ Known (please state below) ○ Unsure

**Q8: Are there any other commissioning services involved?**

○ Yes (please state below) ○ No ○ Unsure

**Q9: How is the catchment areas for your service defined (e.g. by local authority area, CCG area, postcodes, GP practice lists)?**

○ Known (please state below) ○ Unsure

**Q10: What is the catchment area for the local CRHTT where you work?**

○ Known (please state below) ○ Unsure

**Q11: What is the size of the catchment area population?**

○ Known (please state below) ○ Unsure

#### Local acute care system and pathways

**Q12: Is there a written or graphic description of the local mental health acute care pathway and the services involved?**

○ Yes ○ No ○ Unsure

#### Local acute care system and pathways (cont.)

*Note: if you have answered/chosen item [2, 3] in question 12, skip the following question*

**Q13: Can you share this with us?**

- ◯ Yes (please email this to one of our researchers using details at the bottom of this page or upload the image by clicking on the icon next to the text box)
- ◯ No
- ◯ Unsure

#### Local acute care system and pathways (cont.)

**Q14: Is there a first response or crisis assessment service in your area, which provides assessment only and is separate to any home treatment team?A service which can see service users face-to-face and which is: separate from local crisis home treatment team services and A&E psychiatric Liaison teams; will accept referrals for people in mental health crisis; and can see service users face-to-face for initial assessment and referral to appropriate support.**

◯ Yes ◯ No ◯ Unsure

**Q15: Is there a telephone crisis line in your area?**

**A telephone service where: staff can speak to referrers or people in crisis; signpost or refer on to appropriate support; but do not see people face-to-face for assessment.**

◯ Yes ◯ No ◯ Unsure

**Q16: Is there a Crisis Resolution and Home Treatment Team (CRHTT) in your area?**

**A service which: provides intensive home treatment for service users for a few days or weeks, to help resolve a crisis and avoid hospital admission; and which may also provide a first response and initial assessment for people in mental health crisis, where this is not done by a separate crisis assessment service.**

◯ Yes ◯ No ◯ Unsure

#### Crisis assessment services

*A service which can see service users face-to-face and which is: separate from local crisis home treatment team services and A&E Psychiatric Liaison teams; will accept referrals for people in mental health crisis; and can see service users face-to-face for initial assessment and referral to appropriate support.*

*Note: if you have answered/chosen item [2, 3] in question 14, skip the following question*

**Q17: What is the name of the first response or crisis assessment service?**

*Note: if you have answered/chosen item [2, 3] in question 14, skip the following question*

**Q18: Does this first response or crisis assessment service work with any CRHTTs?**

- ◯ Yes (please state which CRHTT(s) below)
- ◯ No
- ◯ Unsure

*Note: if you have answered/chosen item [2, 3] in question 14, skip the following question*

**Q19: Is this crisis assessment service designed to act as a single point of access to all NHS crisis services within its local catchment area?**

- ◯ Yes
- ◯ No (please state below which crisis services this service acts as a single point of access for)
- ◯ Unsure

*Note: if you have answered/chosen item [2, 3] in question 14, skip the following question*

**Q20: What is the lower age limit for people who can be assessed by this service? Please enter 0 if no lower limit or 999 if unsure.**

*Note: if you have answered/chosen item [2, 3] in question 14, skip the following question*

**Q21: What is the upper age limit? Please enter 888 if no upper limit or 999 if unsure.**

*Note: if you have answered/chosen item [2, 3] in question 14, skip the following question*

**Q22: Is it a 24 hour service?**

- ◯ Yes
- ◯ No (please state the opening hours below)
- ◯ Unsure

*Note: if you have answered/chosen item [2, 3] in question 14, skip the following question*

**Q23: Who can refer to this service? Select all that can.**

- ☐ Any member of the public in a mental health crisis or their family, even if not known to services
- ☐ People known to mental health services or their family
- ☐ Non-NHS staff who work in mental health support roles (e.g. voluntary sector workers)
- ☐ NHS111 staff
- ☐ GPs
- ☐ NHS community mental health services
- ☐ Inpatient psychiatric services
- ☐ A&E or psychiatric liaison team staff
- ☐ Police
- ☐ Ambulance workers or paramedics
- ☐ Other (please describe below)
- ☐ Unsure

Please provide any further information below which will help us understand the referral pathways into the first response / crisis assessment service.

*Note: if you have answered/chosen item [2, 3] in question 14, skip the following question*

**Q24: Is there a time limit for how long the crisis assessment service can support someone or hold responsibility for supporting someone in crisis?**

- ◯ Yes (please state the limit in hours below and/or add further explanation)
- ◯ No (please add further explanation below if appropriate)
- ◯ Unsure

*Note: if you have answered/chosen item [2, 3] in question 14, skip the following question*

**Q25: Can the crisis assessment service decide whether someone is suitable to be taken on for CRHTT intensive home treatment care without reassessment from your CRHTT?**

◯ Yes ◯ No ◯ Unsure

*Note: if you have answered/chosen item [2, 3] in question 14, skip the following question*

**Q26: Is there any shared staffing between the crisis assessment service and any other local crisis service?**

- ◯ Yes (please state below which teams)
- ◯ No
- ◯ Unsure

*Note: if you have answered/chosen item [2, 3] in question 14, skip the following question*

**Q27: Is there any joint management at team-level between the crisis assessment service and any other local crisis service?**

- ◯ Yes (please state below which teams and describe the arrangement briefly)
- ◯ No
- ◯ Unsure

### Crisis phone line

*A telephone service where: staff can speak to referrers or people in crisis; signpost or refer on to appropriate support; but do not see people face-to-face for assessment.*

*Note: if you have answered/chosen item [2, 3] in question 15, skip the following question*

**Q28: What is the name of the crisis phone line?**

*Note: if you have answered/chosen item [2, 3] in question 15, skip the following question*

**Q29: What type of organisation provides the service?**

- ◯ NHS
- ◯ Local authority
- ◯ Voluntary sector
- ◯ Unsure

*Note: if you have answered/chosen item [2, 3] in question 15, skip the following question*

**Q30: Is it managed as part of another service (e.g. the CRHTT; the crisis assessment service)?**

- ◯ Yes (please state below which service)
- ◯ No
- ◯ Unsure

*Note: if you have answered/chosen item [2, 3] in question 15, skip the following question*

**Q31: Is it a 24 hour service?**

- ◯ Yes
- ◯ No (please state what the opening hours are below)
- ◯ Unsure

*Note: if you have answered/chosen item [2, 3] in question 15, skip the following question*

**Q32: Who can access this service directly? Please select all that apply**

- ☐ Any member of the public in a mental health crisis or their family, even if not known to services
- ☐ People known to mental health services or their family
- ☐ Non-NHS staff who work in mental health support roles (e.g. voluntary sector workers)
- ☐ NHS111 staff
- ☐ GPs
- ☐ NHS community mental health services
- ☐ Inpatient psychiatric services
- ☐ A&E or psychiatric liaison team staff
- ☐ Police
- ☐ Ambulance workers or paramedics
- ☐ Other (please describe below)
- ☐ Unsure

*Note: if you have answered/chosen item [2, 3] in question 15, skip the following question*

**Q33: Is it just for people living in the catchment area of your service / Trust / local authority?**

- ○ Yes
- ○ No (please describe briefly below)
- ○ Unsure

*Note: if you have answered/chosen item [2, 3] in question 15, skip the following question*

**Q34: Can the crisis line service decide whether someone is suitable to be taken on for CRHTT intensive home treatment care without reassessment from your CRHTT?**

○ Yes ○ No ○ Unsure

*Note: if you have answered/chosen item [2, 3] in question 15, skip the following question*

**Q35: Are there any other local crisis services which the crisis line refer to without reference to the CRHTT (e.g. crisis houses)? Please state all which apply. Please enter None if none.**

- ◯ Yes (please state below which services)
- ◯ No
- ◯ Unsure

*Note: if you have answered/chosen item [2, 3] in question 15, skip the following question*

**Q36: Can the crisis line admit people to hospital without reference to the CRHTT / crisis assessment team?**

- ◯ Yes
- ◯ No
- ◯ Unsure

*Note: if you have answered/chosen item [2, 3] in question 15, skip the following question*

**Q37: Is there any shared staffing between the crisis line and any other local crisis service?**

- ◯ Yes (please state which teams below)
- ◯ No
- ◯ Unsure

*Note: if you have answered/chosen item [2, 3] in question 15, skip the following question*

**Q38: Is there any joint management at team-level between the crisis line and any other local crisis service?**

- ◯ Yes (please state below which teams and describe the arrangement briefly)
- ◯ No
- ◯ Unsure

#### CRHTT services

*Please tell us here about your local Crisis Resolution and Home Treatment Team which provides short-term intensive home treatment to people in mental health crisis. (The CRHTT may also provide initial assessment of referrals for people in mental health crisis, if there is not a separate crisis assessment / first response service.)*

*Note: if you have answered/chosen item [2, 3] in question 16, skip the following question*

**Q39: What is the name of the CRHTT?**

*Note: if you have answered/chosen item [2, 3] in question 16, skip the following question*

**Q40: Does the CRHTT serve Children and Young People (CYP) aged up to 18 when in crisis?**

○ Yes ○ No ○ Unsure

*Note: if you have answered/chosen item [2, 3] in question 16, skip the following question*

**Q41: Does the CRHTT serve working age adults aged between 18 and 65 when in crisis?**

○ Yes ○ No ○ Unsure

*Note: if you have answered/chosen item [2, 3] in question 16, skip the following question*

**Q42: Does the CRHTT serve older age adults aged 65 and up when in crisis?**

○ Yes ○ No ○ Unsure

*Note: if you have answered/chosen item [2, 3] in question 16, skip the following question*

**Q43: Does the CRHTT serve people with dementia when in crisis?**

○ Yes ○ No ○ Unsure

#### CRHTT services (cont.)

*Note: if you have answered/chosen item [1] in question 40, skip the following question*
*Note: if you have answered/chosen item [2, 3] in question 16, skip the following question*

**Q44: Is there a separate CRHTT service in your area for children and young people?**

- ◯ Yes (please state name of service below)
- ◯ No
- ◯ Unsure

#### CRHTT services (cont.)

*Note: if you have answered/chosen item [1] in question 42 AND answered/chosen item [1] in question 43, skip the following question*
*Note: if you have answered/chosen item [2, 3] in question 16, skip the following question*

**Q45: Is there a separate CRHTT service in your area for older adults and/or people with dementia?**

- ◯ Yes, there is a separate older adults crisis service (please state name of service below)
- ◯ Yes, there is a separate dementia crisis service (please state name of service below)
- ◯ No
- ◯ Unsure

#### CRHTT services (cont.)

*In this section, please tell us more about the CRHTT for adults in your area (please don't answer here about any separate CRHTT for older adults, or for children and young people).*

*Note: if you have answered/chosen item [2, 3] in question 16, skip the following question*

**Q46: On weekdays, during which hours does the CRHTT provide home visits?**

- ◯ 24hr
- ◯ Day only (e.g. 8am – 9pm)
- ◯ Night only (e.g. 9pm – 8am)
- ◯ Other (please detail below)
- ◯ Unsure

*Note: if you have answered/chosen item [2, 3] in question 16, skip the following question*

**Q47: On weekdays, during which hours does the CRHTT see patients on NHS premises?**

- ◯ 24hr
- ◯ Day only (e.g. 8am – 9pm)
- ◯ Night only (e.g. 9pm – 8am)
- ◯ Other (please detail below)
- ◯ Unsure

*Note: if you have answered/chosen item [2, 3] in question 16, skip the following question*

**Q48: On weekdays, during which hours does the CRHTT provide phone call support?**

- ◯ 24hr
- ◯ Day only (e.g. 8am – 9pm)
- ◯ Night only (e.g. 9pm – 8am)
- ◯ Other (please detail below)
- ◯ Unsure

*Note: if you have answered/chosen item [2, 3] in question 16, skip the following question*

**Q49: On weekdays, during which hours does the CRHTT provide no service (i.e. the service is closed)?**

- ◯ The service is 24hr (i.e. there are no hours during which there is no service provided)
- ◯ Day only (e.g. 8am – 9pm)
- ◯ Night only (e.g. 9pm – 8am)
- ◯ Other (please detail below)
- ◯ Unsure

*Note: if you have answered/chosen item [2, 3] in question 16, skip the following question*

**Q50: Is service provision different at weekends?**

- ◯ Yes
- ◯ No
- ◯ Unsure

#### CRHTT services (cont.)

*Note: if you have answered/chosen item [2, 3] in question 50, skip the following question*
*Note: if you have answered/chosen item [2, 3] in question 16, skip the following question*

**Q51: On weekends, during which hours does the CRHTT provide home visits?**

- ◯ 24hr
- ◯ Day only (e.g. 8am – 9pm)
- ◯ Night only (e.g. 9pm – 8am)
- ◯ Other (please detail below)
- ◯ Unsure

*Note: if you have answered/chosen item [2, 3] in question 50, skip the following question*
*Note: if you have answered/chosen item [2, 3] in question 16, skip the following question*

**Q52: On weekends, during which hours does the CRHTT see patients on NHS premises?**

- ◯ 24hr
- ◯ Day only (e.g. 8am – 9pm)
- ◯ Night only (e.g. 9pm – 8am)
- ◯ Other (please detail below)
- ◯ Unsure

*Note: if you have answered/chosen item [2, 3] in question 50, skip the following question*
*Note: if you have answered/chosen item [2, 3] in question 16, skip the following question*

**Q53: On weekends, during which hours does the CRHTT provide phone call support?**

- ◯ 24hr
- ◯ Day only (e.g. 8am – 9pm)
- ◯ Night only (e.g. 9pm – 8am)
- ◯ Other (please detail below)
- ◯ Unsure

*Note: if you have answered/chosen item [2, 3] in question 50, skip the following question*
*Note: if you have answered/chosen item [2, 3] in question 16, skip the following question*

**Q54: On weekends, during which hours does the CRHTT provide no service (i.e. the service is closed)?**

◯ The service is 24hr (i.e. there are no hours during which there is no service provided)
◯ Day only (e.g. 8am – 9pm)
◯ Night only (e.g. 9pm – 8am)
◯ Other (please detail below)
◯ Unsure

*Note: if you have answered/chosen item [2, 3] in question 14, skip the following question*
*Note: if you have answered/chosen item [2, 3] in question 16, skip the following question*

**Q55: Do all referrals to the CRHTT have to come through a separate crisis assessment service (including referrals from within the Trust)?**

◯ Yes ◯ No ◯ Unsure

### CRHTT services (cont.)

*Note: if you have answered/chosen item [1] in question 55, skip the following question*
*Note: if you have answered/chosen item [2, 3] in question 16, skip the following question*

**Q56: Which of the following does the CRHTT accept direct referrals from? Please select all that apply.**

☐ Any member of the public in a mental health crisis or their family, even if not known to services
☐ People known to mental health services or their family
☐ Non-NHS staff who work in mental health support roles (e.g. voluntary sector workers)
☐ NHS111 staff
☐ GPs
☐ NHS community mental health services
☐ Inpatient psychiatric services
☐ A&E or psychiatric liaison team staff
☐ Police
☐ Ambulance workers or paramedics
☐ Other (please describe below)
☐ Unsure

Please provide any further information below which will help us understand the referral pathways into the CRHTT service.

*Note: if you have answered/chosen item [2, 3] in question 16, skip the following question*

**Q57: Does the CRHTT have a response time target to see patients in person for assessment following a referral?**

- ◯ Yes
- ◯ No
- ◯ Unsure

### CRHTT response time target

*Note: if you have answered/chosen item [2, 3] in question 57, skip the following question*
*Note: if you have answered/chosen item [2, 3] in question 16, skip the following question*

**Q58: Who does this response time target apply to?**

- ◯ All accepted referrals
- ◯ Only referrals triaged as urgent
- ◯ Unsure

*Note: if you have answered/chosen item [2, 3] in question 57, skip the following question*
*Note: if you have answered/chosen item [2, 3] in question 16, skip the following question*

**Q59: What is the response time target? Please enter 999 if unknown.**

[ ] hours

*Note: if you have answered/chosen item [2, 3] in question 57, skip the following question*
*Note: if you have answered/chosen item [2, 3] in question 16, skip the following question*

**Q60: Do you know how often this response time target is met?**

- ◯ Yes, I know how often it is met (please state % below)
- ◯ No, but I can provide an estimate (please state % below)
- ◯ No

### Inpatient admission gatekeeping functions

**Q61: In what circumstances, if any, do exceptions to crisis assessment team / CRHTT gatekeeping (controlling or limiting inpatient psychiatric admissions) commonly apply? Please select all that apply.**

- ☐ Admissions following a Mental Health Act Assessment
- ☐ When admission is recommended by a trusted assessor (e.g. Psychiatric Liaison or CMHT colleagues)
- ☐ At night time or at weekends when the CRHTT or crisis assessment service is not fully staffed
- ☐ At times when CRHTT or crisis assessment team staff are too busy to assess
- ☐ There are no exceptions
- ☐ Unsure

**Q62: What proportion of patients admitted to acute psychiatric hospital inpatient beds are first assessed in person by the CRHTT or the local crisis assessment team regarding suitability for home treatment (i.e. involving face-to-face meeting with the patient, not just through a phone call or discussion with the referrer)?**

**Please select the answer which best describes arrangements for your team. Please estimate if data is not available.**

- ○ The CRHTT or crisis assessment team assesses at least 95% of patients in person before they are admitted to an acute inpatient bed
- ○ The CRHTT or crisis assessment team assesses at least 80% of patients in person before they are admitted to an acute inpatient bed
- ○ The CRHTT or crisis assessment team assesses at least 50% of patients in person before they are admitted to an acute inpatient bed
- ○ The CRHTT or crisis assessment team assesses fewer than of 50% patients in person before they are admitted to an acute inpatient bed
- ○ Unsure

**Q63: Is this figure based on:**

○ Locally collected data ○ Your best estimate ○ Not applicable

**Q64: Is there a local protocol which specifies gatekeeping arrangements for admission to inpatient wards?**

○ Yes ○ No ○ Unsure

### Inpatient admission gatekeeping functions (cont.)

*Note: if you have answered/chosen item [2, 3] in question 64, skip the following question*

**Q65: Does it specify that gatekeeping functions may be completed by the CRHTT or crisis assessment team through phone discussion?**

◯ Yes ◯ No ◯ Unsure

*Note: if you have answered/chosen item [2, 3] in question 64, skip the following question*

**Q66: Does it specify any “trusted assessors” (e.g. psychiatric liaison or CMHT colleagues) who may admit patients to hospital without involvement from the CRHTT or crisis assessment team?**

◯ Yes ◯ No ◯ Unsure

#### Psychiatric liaison services

**Q67: Is there a 24-hour psychiatric liaison service at Accident and Emergency (AandE) departments in the hospital(s) local to your CRHTT?**

◯ Yes
◯ No (please describe below what hours a psychiatric liaison service is open)
◯ Unsure

**Q68: Is there any shared staffing between the psychiatric liaison service and any other local crisis service?**

◯ Yes (please state which teams below) ◯ No
◯ Unsure

**Q69: How often do staff from the CRHTT or a separate local crisis assessment service attend assessments at AandE with psychiatric liaison when home treatment is being considered?**

◯ Always ◯ Usually ◯ Sometimes ◯ Never ◯ Unsure

**Q70: Is there any joint management at team-level between the psychiatric liaison team and any other local crisis service?**

- ◯ Yes (please state below which teams and describe the arrangement briefly)
- ◯ No
- ◯ Unsure

**Q71: If referral to the local crisis assessment team or CRHTT is not available 24-hours, is there any other local service to which psychiatric liaison can refer people for immediate crisis support?**

- ◯ Yes (please describe below)
- ◯ No
- ◯ Not applicable (referral to crisis assessment team or CRHTT is available 24-hours)
- ◯ Unsure

**Q72: When can psychiatric liaison admit to hospital?**

- ◯ Without CRHTT or crisis assessment service input
- ◯ Following phone discussion with CRHTT / crisis assessment service
- ◯ Only following in person assessment of the patient by CRHTT / crisis assessment service
- ◯ Other (please describe below)
- ◯ Unsure

#### Psychiatric Decision Units and Triage wards

**Q73: Is there a Psychiatric Decision Unit (PDU) in your local area?**

**A PDU is a dedicated area (separate to an A&E department / psychiatric ward / psychiatric liaison team) in which assessment can be conducted and treatment plans developed for patients in mental health crisis who are accessing emergency services. People may typically stay for up to 24-48 hours in a PDU, which may be called a “Clinical Decision Unit” or other in some areas. Please only tell us about services called a PDU here. We will ask about “crisis café” or “Haven” style services later in this survey.**

○ Yes ○ No ○ Unsure

**Q74: Is there a triage ward in your local area?**

**A triage ward is an inpatient psychiatric ward which only accepts admissions for a time-limited period (not more than one week); typically does not people compulsorily admitted on a MHA section; and works closely with local community crisis services to avoid the need to transfer to an acute psychiatric ward.**

○ Yes ○ No ○ Unsure

#### PDU

*Note: if you have answered/chosen item [2, 3] in question 73, skip the following question*

**Q75: What is the name of the PDU?**

*Note: if you have answered/chosen item [2, 3] in question 73, skip the following question*

**Q76: Where is the PDU located?**

○ Psychiatric hospital ○ Acute hospital ○ Other (please specify below)
○ Unsure

*Note: if you have answered/chosen item [2, 3] in question 73, skip the following question*

**Q77: What hours is the PDU open?**

- ◯ 24hr
- ◯ 24hr Monday to Friday and 9am – 5pm Saturday and Sunday
- ◯ 24hr Monday to Friday
- ◯ 9am – 5pm Monday to Sunday
- ◯ 9am – 5pm Monday to Friday
- ◯ Other (please detail below)
- ◯ Unsure

*Note: if you have answered/chosen item [2, 3] in question 73, skip the following question*

**Q78: What is the accommodation on the PDU like?**

- ◯ Single rooms / partitioned area
- ◯ Beds
- ◯ Recliners
- ◯ No overnight accommodation
- ◯ Other (please describe below)
- ◯ Unsure

*Note: if you have answered/chosen item [2, 3] in question 73, skip the following question*

**Q79: How long is the maximum length of stay on the unit? Please enter 999 if unsure.**

________ hours

*Note: if you have answered/chosen item [2, 3] in question 73, skip the following question*

**Q80: Who can refer to the PDU? Please select all that apply.**

- ☐ CRHTT or crisis assessment team
- ☐ Psychiatric liaison service
- ☐ Street triage team
- ☐ Community mental health teams
- ☐ GPs
- ☐ Third/ voluntary sector services
- ☐ Self-referral
- ☐ Other (please specify below)
- ☐ Unsure

Please provide any further information below which will help us understand the referral pathways into the PDU.

*Note: if you have answered/chosen item [2, 3] in question 73, skip the following question*

**Q81: Is the PDU co-located with any other services (e.g. Section 136 Place of Safety)? Please state. Enter 999 if unsure.**

- ◯ Yes (please state which service below)
- ◯ No
- ◯ Unsure

*Note: if you have answered/chosen item [2, 3] in question 73, skip the following question*

**Q82: Is there any shared staffing between the PDU and any other local crisis service?**

- ◯ Yes (please state which services below)
- ◯ No
- ◯ Unsure

*Note: if you have answered/chosen item [2, 3] in question 73, skip the following question*

**Q83: What is the capacity of the PDU i.e. how many people can it admit at any one time? Enter 999 if unsure.**

*Note: if you have answered/chosen item [2, 3] in question 73, skip the following question*

**Q84: How often do staff from the CRHTT or a separate local crisis assessment service attend assessments at the PDU when home treatment is being considered?**

◯ Always ◯ Usually ◯ Sometimes ◯ Never ◯ Unsure

*Note: if you have answered/chosen item [2, 3] in question 73, skip the following question*

**Q85: When can the PDU admit patients to hospital?**

- ◯ Without CRHTT or crisis assessment service input
- ◯ Following phone discussion with CRHTT / crisis assessment service
- ◯ Only following in person assessment of the patient by CRHTT / crisis assessment service
- ◯ Other (please describe below)
- ◯ Unsure

*Note: if you have answered/chosen item [2, 3] in question 73, skip the following question*

**Q86: Is there any joint management at team-level between the PDU and any other local crisis service?**

- ◯ Yes (please state below which teams and describe the arrangement briefly)
- ◯ No
- ◯ Unsure

### Triage ward

*Note: if you have answered/chosen item [2, 3] in question 74, skip the following question*

**Q87: What is the name of the triage ward?**

*Note: if you have answered/chosen item [2, 3] in question 74, skip the following question*

**Q88: How many beds does it have? Please enter 999 if unsure.**

[ ]

*Note: if you have answered/chosen item [2, 3] in question 74, skip the following question*

**Q89: How long is the maximum stay on the triage ward? Please enter 999 if unsure.**

[ ] hours

*Note: if you have answered/chosen item [2, 3] in question 74, skip the following question*

**Q90: Who can refer to the triage ward? Please select all that apply.**

- ☐ CRHTT or crisis assessment team
- ☐ Psychiatric liaison service
- ☐ Street triage team
- ☐ Community mental health teams
- ☐ GPs
- ☐ Third / voluntary sector services
- ☐ Self-referral
- ☐ Other (please specify below)
- ☐ Unsure

Please provide any further information below which will help us understand the referral pathways into the triage ward.

*Note: if you have answered/chosen item [2, 3] in question 74, skip the following question*

**Q91: Is the triage ward co-located with any other services (e.g. Section 136 Place of Safety)? Please state. Enter 999 if unsure.**

*Note: if you have answered/chosen item [2, 3] in question 74, skip the following question*

**Q92: Is there any shared staffing between the triage ward and any other local crisis service?**

- ○ Yes (please state which services below)
- ○ No
- ○ Unsure

*Note: if you have answered/chosen item [2, 3] in question 74, skip the following question*

**Q93: How often do staff from the CRHTT or a separate local crisis assessment service attend assessments at the triage ward when home treatment is being considered?**

- ○ Always
- ○ Usually
- ○ Sometimes
- ○ Never
- ○ Unsure

*Note: if you have answered/chosen item [2, 3] in question 74, skip the following question*

**Q94: When can the triage ward admit a patient to another acute psychiatric ward?**

- ○ Without CRHTT or crisis assessment service input
- ○ Following phone discussion with CRHTT / crisis assessment service
- ○ Only following in person assessment of the patient by CRHTT / crisis assessment service
- ○ Other (please describe below)
- ○ Unsure

*Note: if you have answered/chosen item [2, 3] in question 74, skip the following question*

**Q95: Is there any joint management at team-level between the triage and any other local crisis service?**

- ○ Yes (please state below which teams and describe the arrangement briefly)
- ○ No
- ○ Unsure

#### Crisis Houses

*Residential services not classified as hospitals for people in mental health crisis. Please do not tell us about respite or rehabilitation services here, only crisis houses for people in a severe mental health crisis.*

**Q96: Are there any non-hospital, residential crisis services / crisis beds (crisis house) in your area?**

○ Yes ○ No ○ Unsure

#### Crisis house one

*Note: if you have answered/chosen item [2, 3] in question 96, skip the following question*

**Q97: What is the name of the service?Please only name one service. We will later ask you if there are any other services of this type in your area.**

*Note: if you have answered/chosen item [2, 3] in question 96, skip the following question*

**Q98: What type of organisation provides the service?**

○ NHS ○ Local authority ○ Voluntary sector ○ Unsure

*Note: if you have answered/chosen item [2, 3] in question 96, skip the following question*

**Q99: Referral pathways (please select all that apply):**

☐ The CRHTT and/or crisis assessment service can refer to this service
☐ The CRHTT and/or crisis assessment service have exclusive referral rights
☐ People in crisis can self-refer to this service
☐ Unsure

Please provide any further information below which will help us understand the referral pathways into the crisis house.

*Note: if you have answered/chosen item [2, 3] in question 96, skip the following question*

**Q100: What is the lower age limit for people who can use the crisis house? Please enter 0 if no minimum or 999 if unsure.**

*Note: if you have answered/chosen item [2, 3] in question 96, skip the following question*

**Q101: What is the upper age limit? Please enter 888 if no minimum or 999 if unsure.**

*Note: if you have answered/chosen item [2, 3] in question 96, skip the following question*

**Q102: Is this crisis house for any other specific patient group (e.g. a women's crisis house, for specific diagnoses, risk thresholds etc.)?**

- ○ Yes (please state which groups below)
- ○ No
- ○ Unsure

*Note: if you have answered/chosen item [2, 3] in question 96, skip the following question*

**Q103: Can the CRHTT or crisis assessment service access a place at this service for a patient when needed?**

- ○ Always
- ○ Usually
- ○ Sometimes
- ○ Rarely
- ○ Unsure

*Note: if you have answered/chosen item [2, 3] in question 96, skip the following question*

**Q104: How quickly can admission to this service typically be arranged following initial referral?**

- ○ In less than four hours
- ○ Within 24 hours
- ○ Within 72 hours
- ○ Within one week
- ○ Longer than one week
- ○ Unsure

*Note: if you have answered/chosen item [2, 3] in question 96, skip the following question*

**Q105: What is the maximum length of stay at this service? Please enter 999 if unsure.**

days

*Note: if you have answered/chosen item [2, 3] in question 96, skip the following question*

**Q106: What is a typical length of stay at this service? Please enter 999 if unsure.**

[ ] days

*Note: if you have answered/chosen item [2, 3] in question 96, skip the following question*

**Q107: Is this crisis house service user-led?**

○ Yes ○ No ○ Unsure

*Note: if you have answered/chosen item [2, 3] in question 96, skip the following question*

**Q108: How many of the crisis house staff are clinicians with mental health professional qualifications (e.g. nurses or social workers)?**

○ All of the staff ○ Most of the staff ○ Some of the staff ○ None of the staff ○ Unsure

*Note: if you have answered/chosen item [2, 3] in question 96, skip the following question*

**Q109: Is there any shared staffing between this crisis house and any other local crisis service?**

○ Yes (please state below which teams and describe the arrangement briefly)
○ No
○ Unsure

*Note: if you have answered/chosen item [2, 3] in question 96, skip the following question*

**Q110: Is there any joint management at team-level between this crisis house and any other local crisis service?**

○ Yes (please state below which teams and describe the arrangement briefly)
○ No
○ Unsure

*Note: if you have answered/chosen item [2, 3] in question 96, skip the following question*

**Q111: Are there any other residential crisis services in your area?**

○ Yes ○ No ○ Unsure

### Crisis house two

*Note: if you have answered/chosen item [2, 3] in question 96, skip the following question*
*Note: if you have answered/chosen item [2, 3] in question 111, skip the following question*

**Q112: What is the name of the service?**

*Note: if you have answered/chosen item [2, 3] in question 96, skip the following question*
*Note: if you have answered/chosen item [2, 3] in question 111, skip the following question*

**Q113: What type of organisation provides the service?**

○ NHS ○ Local authority ○ Voluntary sector ○ Unsure

*Note: if you have answered/chosen item [2, 3] in question 96, skip the following question*
*Note: if you have answered/chosen item [2, 3] in question 111, skip the following question*

**Q114: Referral pathways (please select all that apply):**

☐ The CRHTT and/or crisis assessment service can refer to this service
☐ The CRHTT and/or crisis assessment service have exclusive referral rights
☐ People in crisis can self-refer to this service
☐ Unsure

Please provide any further information below which will help us understand the referral pathways into the crisis house.

*Note: if you have answered/chosen item [2, 3] in question 96, skip the following question*
*Note: if you have answered/chosen item [2, 3] in question 111, skip the following question*

**Q115: What is the lower age limit for people who can use the crisis house? Please enter 0 if no minimum or 999 if unsure.**

*Note: if you have answered/chosen item [2, 3] in question 96, skip the following question*
*Note: if you have answered/chosen item [2, 3] in question 111, skip the following question*

**Q116: What is the upper age limit? Please enter 888 if no minimum or 999 if unsure.**

*Note: if you have answered/chosen item [2, 3] in question 96, skip the following question*
*Note: if you have answered/chosen item [2, 3] in question 111, skip the following question*

**Q117: Is this crisis house for any other specific patient group (e.g. a women's crisis house, for specific diagnoses, risk thresholds etc.)?**

◯ Yes (please state which groups below) ◯ No
◯ Unsure

*Note: if you have answered/chosen item [2, 3] in question 96, skip the following question*
*Note: if you have answered/chosen item [2, 3] in question 111, skip the following question*

**Q118: Can the CRHTT or crisis assessment service access a place at this service for a patient when needed?**

◯ Always ◯ Usually ◯ Sometimes ◯ Rarely ◯ Unsure

*Note: if you have answered/chosen item [2, 3] in question 96, skip the following question*
*Note: if you have answered/chosen item [2, 3] in question 111, skip the following question*

**Q119: How quickly can admission to this service typically be arranged following initial referral?**

○ In less than four hours ○ Within 24 hours ○ Within 72 hours ○ Within one week
○ Longer than one week ○ Unsure

*Note: if you have answered/chosen item [2, 3] in question 96, skip the following question*
*Note: if you have answered/chosen item [2, 3] in question 111, skip the following question*

**Q120: What is the maximum length of stay at this service? Please enter 999 if unsure.**

[ ] days

*Note: if you have answered/chosen item [2, 3] in question 96, skip the following question*
*Note: if you have answered/chosen item [2, 3] in question 111, skip the following question*

**Q121: What is a typical length of stay at this service? Please enter 999 if unsure.**

[ ] days

*Note: if you have answered/chosen item [2, 3] in question 96, skip the following question*
*Note: if you have answered/chosen item [2, 3] in question 111, skip the following question*

**Q122: Is this crisis house service user-led?**

○ Yes ○ No ○ Unsure

*Note: if you have answered/chosen item [2, 3] in question 96, skip the following question*
*Note: if you have answered/chosen item [2, 3] in question 111, skip the following question*

**Q123: How many of the crisis house staff are clinicians with mental health professional qualifications (e.g. nurses or social workers)?**

○ All of the staff ○ Most of the staff ○ Some of the staff ○ None of the staff ○ Unsure

*Note: if you have answered/chosen item [2, 3] in question 96, skip the following question*
*Note: if you have answered/chosen item [2, 3] in question 111, skip the following question*

**Q124: Is there any shared staffing between this crisis house and any other local crisis service?**

○ Yes (please state below which teams and describe the arrangement briefly)
○ No
○ Unsure

*Note: if you have answered/chosen item [2, 3] in question 96, skip the following question*
*Note: if you have answered/chosen item [2, 3] in question 111, skip the following question*

**Q125: Is there any joint management at team-level between this crisis house and any other local crisis service?**

- ◯ Yes (please state below which teams and describe the arrangement briefly)
- ◯ No
- ◯ Unsure

*Note: if you have answered/chosen item [2, 3] in question 96, skip the following question*
*Note: if you have answered/chosen item [2, 3] in question 111, skip the following question*

**Q126: Are there any other residential crisis services in your area?**

◯ Yes ◯ No ◯ Unsure

#### Crisis house three

*Note: if you have answered/chosen item [2, 3] in question 96, skip the following question*
*Note: if you have answered/chosen item [2, 3] in question 111, skip the following question*
*Note: if you have answered/chosen item [2, 3] in question 126, skip the following question*

**Q127: What is the name of the service?**

*Note: if you have answered/chosen item [2, 3] in question 96, skip the following question*
*Note: if you have answered/chosen item [2, 3] in question 111, skip the following question*
*Note: if you have answered/chosen item [2, 3] in question 126, skip the following question*

**Q128: What type of organisation provides the service?**

◯ NHS ◯ Local authority ◯ Voluntary sector ◯ Unsure

*Note: if you have answered/chosen item [2, 3] in question 96, skip the following question*
*Note: if you have answered/chosen item [2, 3] in question 111, skip the following question*
*Note: if you have answered/chosen item [2, 3] in question 126, skip the following question*

**Q129: Referral pathways (please select all that apply):**

☐ The CRHTT and/or crisis assessment service can refer to this service
☐ The CRHTT and/or crisis assessment service have exclusive referral rights
☐ People in crisis can self-refer to this service
☐ Unsure

Please provide any further information below which will help us understand the referral pathways into the crisis house.

*Note: if you have answered/chosen item [2, 3] in question 96, skip the following question*
*Note: if you have answered/chosen item [2, 3] in question 111, skip the following question*
*Note: if you have answered/chosen item [2, 3] in question 126, skip the following question*

**Q130: What is the lower age limit for people who can use the crisis house? Please enter 0 if no minimum or 999 if unsure.**

*Note: if you have answered/chosen item [2, 3] in question 96, skip the following question*
*Note: if you have answered/chosen item [2, 3] in question 111, skip the following question*
*Note: if you have answered/chosen item [2, 3] in question 126, skip the following question*

**Q131: What is the upper age limit? Please enter 888 if no minimum or 999 if unsure.**

*Note: if you have answered/chosen item [2, 3] in question 96, skip the following question*
*Note: if you have answered/chosen item [2, 3] in question 111, skip the following question*
*Note: if you have answered/chosen item [2, 3] in question 126, skip the following question*

**Q132: Is this crisis house for any other specific patient group (e.g. a women's crisis house, for specific diagnoses, risk thresholds etc.)?**

◯ Yes (please state which groups below) ◯ No
◯ Unsure

*Note: if you have answered/chosen item [2, 3] in question 96, skip the following question*
*Note: if you have answered/chosen item [2, 3] in question 111, skip the following question*
*Note: if you have answered/chosen item [2, 3] in question 126, skip the following question*

**Q133: Can the CRHTT or crisis assessment service access a place at this service for a patient when needed?**

○ Always ○ Usually ○ Sometimes ○ Rarely ○ Unsure

*Note: if you have answered/chosen item [2, 3] in question 96, skip the following question*
*Note: if you have answered/chosen item [2, 3] in question 111, skip the following question*
*Note: if you have answered/chosen item [2, 3] in question 126, skip the following question*

**Q134: How quickly can admission to this service typically be arranged following initial referral?**

○ In less than four hours ○ Within 24 hours ○ Within 72 hours ○ Within one week
○ Longer than one week ○ Unsure

*Note: if you have answered/chosen item [2, 3] in question 96, skip the following question*
*Note: if you have answered/chosen item [2, 3] in question 111, skip the following question*
*Note: if you have answered/chosen item [2, 3] in question 126, skip the following question*

**Q135: What is the maximum length of stay at this service? Please enter 999 if unsure.**

[ ] days

*Note: if you have answered/chosen item [2, 3] in question 96, skip the following question*
*Note: if you have answered/chosen item [2, 3] in question 111, skip the following question*
*Note: if you have answered/chosen item [2, 3] in question 126, skip the following question*

**Q136: What is a typical length of stay at this service? Please enter 999 if unsure.**

[ ] days

*Note: if you have answered/chosen item [2, 3] in question 96, skip the following question*
*Note: if you have answered/chosen item [2, 3] in question 111, skip the following question*
*Note: if you have answered/chosen item [2, 3] in question 126, skip the following question*

**Q137: Is this crisis house service user-led?**

○ Yes ○ No ○ Unsure

*Note: if you have answered/chosen item [2, 3] in question 96, skip the following question*
*Note: if you have answered/chosen item [2, 3] in question 111, skip the following question*
*Note: if you have answered/chosen item [2, 3] in question 126, skip the following question*

**Q138: How many of the crisis house staff are clinicians with mental health professional qualifications (e.g. nurses or social workers)?**

○ All of the staff ○ Most of the staff ○ Some of the staff ○ None of the staff ○ Unsure

*Note: if you have answered/chosen item [2, 3] in question 96, skip the following question*
*Note: if you have answered/chosen item [2, 3] in question 111, skip the following question*
*Note: if you have answered/chosen item [2, 3] in question 126, skip the following question*

**Q139: Is there any shared staffing between this crisis house and any other local crisis service?**

- ◯ Yes (please state below which teams and describe the arrangement briefly)
- ◯ No
- ◯ Unsure

*Note: if you have answered/chosen item [2, 3] in question 96, skip the following question*
*Note: if you have answered/chosen item [2, 3] in question 111, skip the following question*
*Note: if you have answered/chosen item [2, 3] in question 126, skip the following question*

**Q140: Is there any joint management at team-level between this crisis house and any other local crisis service?**

- ◯ Yes (please state below which teams and describe the arrangement briefly)
- ◯ No
- ◯ Unsure

*Note: if you have answered/chosen item [2, 3] in question 96, skip the following question*
*Note: if you have answered/chosen item [2, 3] in question 111, skip the following question*
*Note: if you have answered/chosen item [2, 3] in question 126, skip the following question*

**Q141: Are there any other residential crisis services (crisis houses) in your area?**

◯ Yes ◯ No ◯ Unsure

#### All other crisis houses

*Note: if you have answered/chosen item [2, 3] in question 96, skip the following question*
*Note: if you have answered/chosen item [2, 3] in question 111, skip the following question*
*Note: if you have answered/chosen item [2, 3] in question 126, skip the following question*
*Note: if you have answered/chosen item [2, 3] in question 141, skip the following question*

**Q142: Please list all other residential crisis services (crisis houses):**

#### Acute day unit

**Q143: Is there an Acute Day Unit in your area?**

**An acute day unit is a non-residential day hospital / crisis day service providing activity and therapeutic groups, for people in serious mental health crisis. Please do not tell us about longer-term recovery or rehabilitation day services here.**

○ Yes ○ No ○ Unsure

#### Acute day unit one

*Note: if you have answered/chosen item [2, 3] in question 143, skip the following question*

**Q144: What is the name of the service?Please only name one service. We will later ask you if there are any other services of this type in your area.**

*Note: if you have answered/chosen item [2, 3] in question 143, skip the following question*

**Q145: What type of organisation provides the service?**

○ NHS ○ Local authority ○ Voluntary sector ○ Unsure

*Note: if you have answered/chosen item [2, 3] in question 143, skip the following question*

**Q146: Referral pathways (please select all that apply):**

☐ The CRHTT and/or crisis assessment service can refer to this service
☐ The CRHTT and/or crisis assessment service have exclusive referral rights
☐ People in crisis can self-refer to this service
☐ Unsure

Please provide any further information below which will help us understand the referral pathways into the acute day unit.

*Note: if you have answered/chosen item [2, 3] in question 143, skip the following question*

**Q147: What is the lower age limit for people who can use the acute day unit? Please enter 0 if no minimum or 999 if unsure.**

*Note: if you have answered/chosen item [2, 3] in question 143, skip the following question*

**Q148: What is the upper age limit? Please enter 888 if no minimum or 999 if unsure.**

*Note: if you have answered/chosen item [2, 3] in question 143, skip the following question*

**Q149: Is this acute day unit for any other specific patient group (e.g. specific diagnosis or demographic group, risk thresholds etc.)?**

- ◯ Yes (please state which groups below)
- ◯ No
- ◯ Unsure

*Note: if you have answered/chosen item [2, 3] in question 143, skip the following question*

**Q150: Can the CRHTT or crisis assessment service access a place at this service for a patient when needed?**

- ◯ Always
- ◯ Usually
- ◯ Sometimes
- ◯ Rarely
- ◯ Unsure

*Note: if you have answered/chosen item [2, 3] in question 143, skip the following question*

**Q151: How quickly can admission to this service typically be arranged following initial referral?**

- ◯ In less than four hours
- ◯ Within 24 hours
- ◯ Within 72 hours
- ◯ Within one week
- ◯ Longer than one week
- ◯ Unsure

*Note: if you have answered/chosen item [2, 3] in question 143, skip the following question*

**Q152: What is the maximum period of time a patient can use this service? Please enter 999 if unsure.**

[ ] days

*Note: if you have answered/chosen item [2, 3] in question 143, skip the following question*

**Q153: What is the typical period of time a patient can use this service? Please enter 999 if unsure.**

[ ] days

*Note: if you have answered/chosen item [2, 3] in question 143, skip the following question*

**Q154: Is this acute day unit service user-led?**

○ Yes ○ No ○ Unsure

*Note: if you have answered/chosen item [2, 3] in question 143, skip the following question*

**Q155: How many of the acute day unit staff are clinicians with mental health professional qualifications (e.g. nurses or social workers)?**

○ All of the staff ○ Most of the staff ○ Some of the staff ○ None of the staff ○ Unsure

*Note: if you have answered/chosen item [2, 3] in question 143, skip the following question*

**Q156: Is there any shared staffing between this acute day unit and any other local crisis service?**

○ Yes (please state below which teams and describe the arrangement briefly)
○ No
○ Unsure

*Note: if you have answered/chosen item [2, 3] in question 143, skip the following question*

**Q157: Is there any joint management at team-level between this acute day unit and any other local crisis service?**

○ Yes (please state below which teams and describe the arrangement briefly)
○ No
○ Unsure

*Note: if you have answered/chosen item [2, 3] in question 143, skip the following question*

**Q158: Are there any other acute day units in your area?**

◯ Yes ◯ No ◯ Unsure

#### Acute day unit two

*Note: if you have answered/chosen item [2, 3] in question 143, skip the following question*
*Note: if you have answered/chosen item [2, 3] in question 158, skip the following question*

**Q159: What is the name of the service?**

*Note: if you have answered/chosen item [2, 3] in question 143, skip the following question*
*Note: if you have answered/chosen item [2, 3] in question 158, skip the following question*

**Q160: What type of organisation provides the service?**

◯ NHS ◯ Local authority ◯ Voluntary sector ◯ Unsure

*Note: if you have answered/chosen item [2, 3] in question 143, skip the following question*
*Note: if you have answered/chosen item [2, 3] in question 158, skip the following question*

**Q161: Referral pathways (please select all that apply):**

- ☐ The CRHTT and/or crisis assessment service can refer to this service
- ☐ The CRHTT and/or crisis assessment service have exclusive referral rights
- ☐ People in crisis can self-refer to this service
- ☐ Unsure

Please provide any further information below which will help us understand the referral pathways into the acute day unit.

*Note: if you have answered/chosen item [2, 3] in question 143, skip the following question*
*Note: if you have answered/chosen item [2, 3] in question 158, skip the following question*

**Q162: What is the lower age limit for people who can use the acute day unit? Please enter 0 if no minimum or 999 if unsure.**

*Note: if you have answered/chosen item [2, 3] in question 143, skip the following question*
*Note: if you have answered/chosen item [2, 3] in question 158, skip the following question*

**Q163: What is the upper age limit? Please enter 888 if no minimum or 999 if unsure.**

*Note: if you have answered/chosen item [2, 3] in question 143, skip the following question*
*Note: if you have answered/chosen item [2, 3] in question 158, skip the following question*

**Q164: Is this acute day unit for any other specific patient group (e.g. specific diagnosis or demographic group, risk thresholds etc.)?**

○ Yes (please state which groups below) ○ No
○ Unsure

*Note: if you have answered/chosen item [2, 3] in question 143, skip the following question*
*Note: if you have answered/chosen item [2, 3] in question 158, skip the following question*

**Q165: Can the CRHTT or crisis assessment service access a place at this service for a patient when needed?**

○ Always ○ Usually ○ Sometimes ○ Rarely ○ Unsure

*Note: if you have answered/chosen item [2, 3] in question 143, skip the following question*
*Note: if you have answered/chosen item [2, 3] in question 158, skip the following question*

**Q166: How quickly can admission to this service typically be arranged following initial referral?**

○ In less than four hours ○ Within 24 hours ○ Within 72 hours ○ Within one week
○ Longer than one week ○ Unsure

*Note: if you have answered/chosen item [2, 3] in question 143, skip the following question*
*Note: if you have answered/chosen item [2, 3] in question 158, skip the following question*

**Q167: What is the maximum period of time a patient can use this service? Please enter 999 if unsure.**

[ ] days

*Note: if you have answered/chosen item [2, 3] in question 143, skip the following question*
*Note: if you have answered/chosen item [2, 3] in question 158, skip the following question*

**Q168: What is the typical period of time a patient can use this service? Please enter 999 if unsure.**

[ ] days

*Note: if you have answered/chosen item [2, 3] in question 143, skip the following question*
*Note: if you have answered/chosen item [2, 3] in question 158, skip the following question*

**Q169: Is the acute day unit service user-led?**

○ Yes ○ No ○ Unsure

*Note: if you have answered/chosen item [2, 3] in question 143, skip the following question*
*Note: if you have answered/chosen item [2, 3] in question 158, skip the following question*

**Q170: How many of the acute day unit staff are clinicians with mental health professional qualifications (e.g. nurses or social workers)?**

○ All of the staff ○ Most of the staff ○ Some of the staff ○ None of the staff ○ Unsure

*Note: if you have answered/chosen item [2, 3] in question 143, skip the following question*
*Note: if you have answered/chosen item [2, 3] in question 158, skip the following question*

**Q171: Is there any shared staffing between this acute day unit and any other local crisis service?**

○ Yes (please state below which teams and describe the arrangement briefly)
○ No
○ Unsure

*Note: if you have answered/chosen item [2, 3] in question 143, skip the following question*
*Note: if you have answered/chosen item [2, 3] in question 158, skip the following question*

**Q172: Is there any joint management at team-level between this acute day unit and any other local crisis service?**

- ◯ Yes (please state below which teams and describe the arrangement briefly)
- ◯ No
- ◯ Unsure

*Note: if you have answered/chosen item [2, 3] in question 143, skip the following question*
*Note: if you have answered/chosen item [2, 3] in question 158, skip the following question*

**Q173: Are there any other acute day units in your area?**

◯ Yes ◯ No ◯ Unsure

#### Acute day unit three

*Note: if you have answered/chosen item [2, 3] in question 143, skip the following question*
*Note: if you have answered/chosen item [2, 3] in question 158, skip the following question*
*Note: if you have answered/chosen item [2, 3] in question 173, skip the following question*

**Q174: What is the name of the service?**

*Note: if you have answered/chosen item [2, 3] in question 143, skip the following question*
*Note: if you have answered/chosen item [2, 3] in question 158, skip the following question*
*Note: if you have answered/chosen item [2, 3] in question 173, skip the following question*

**Q175: What type of organisation provides the service?**

◯ NHS ◯ Local authority ◯ Voluntary sector ◯ Unsure

*Note: if you have answered/chosen item [2, 3] in question 143, skip the following question*
*Note: if you have answered/chosen item [2, 3] in question 158, skip the following question*
*Note: if you have answered/chosen item [2, 3] in question 173, skip the following question*

**Q176: Referral pathways (please select all that apply):**

- ☐ The CRHTT and/or crisis assessment service can refer to this service
- ☐ The CRHTT and/or crisis assessment service have exclusive referral rights
- ☐ People in crisis can self-refer to this service
- ☐ Unsure

Please provide any further information below which will help us understand the referral pathways into the acute day unit.

*Note: if you have answered/chosen item [2, 3] in question 143, skip the following question*
*Note: if you have answered/chosen item [2, 3] in question 158, skip the following question*
*Note: if you have answered/chosen item [2, 3] in question 173, skip the following question*

**Q177: What is the lower age limit for people who can use the acute day unit? Please enter 0 if no minimum or 999 if unsure.**

*Note: if you have answered/chosen item [2, 3] in question 143, skip the following question*
*Note: if you have answered/chosen item [2, 3] in question 158, skip the following question*
*Note: if you have answered/chosen item [2, 3] in question 173, skip the following question*

**Q178: What is the upper age limit? Please enter 888 if no minimum or 999 if unsure.**

*Note: if you have answered/chosen item [2, 3] in question 143, skip the following question*
*Note: if you have answered/chosen item [2, 3] in question 158, skip the following question*
*Note: if you have answered/chosen item [2, 3] in question 173, skip the following question*

**Q179: Is this acute day unit for any other specific patient group (e.g. specific diagnosis or demographic group, risk thresholds etc.)?**

- ○ Yes (please state which groups below)
- ○ No
- ○ Unsure

*Note: if you have answered/chosen item [2, 3] in question 143, skip the following question*
*Note: if you have answered/chosen item [2, 3] in question 158, skip the following question*
*Note: if you have answered/chosen item [2, 3] in question 173, skip the following question*

**Q180: Can the CRHTT or crisis assessment service access a place at this service for a patient when needed?**

- ◯ Always
- ◯ Usually
- ◯ Sometimes
- ◯ Rarely
- ◯ Unsure

*Note: if you have answered/chosen item [2, 3] in question 143, skip the following question*
*Note: if you have answered/chosen item [2, 3] in question 158, skip the following question*
*Note: if you have answered/chosen item [2, 3] in question 173, skip the following question*

**Q181: How quickly can admission to this service typically be arranged following initial referral?**

- ◯ In less than four hours
- ◯ Within 24 hours
- ◯ Within 72 hours
- ◯ Within one week
- ◯ Longer than one week
- ◯ Unsure

*Note: if you have answered/chosen item [2, 3] in question 143, skip the following question*
*Note: if you have answered/chosen item [2, 3] in question 158, skip the following question*
*Note: if you have answered/chosen item [2, 3] in question 173, skip the following question*

**Q182: What is the maximum period of time a patient can use this service? Please enter 999 if unsure.**

[ ] days

*Note: if you have answered/chosen item [2, 3] in question 143, skip the following question*
*Note: if you have answered/chosen item [2, 3] in question 158, skip the following question*
*Note: if you have answered/chosen item [2, 3] in question 173, skip the following question*

**Q183: What is the typical period of time a patient can use this service? Please enter 999 if unsure.**

[ ] days

*Note: if you have answered/chosen item [2, 3] in question 143, skip the following question*
*Note: if you have answered/chosen item [2, 3] in question 158, skip the following question*
*Note: if you have answered/chosen item [2, 3] in question 173, skip the following question*

**Q184: Is this acute day unit service user-led?**

- ◯ Yes
- ◯ No
- ◯ Unsure

*Note: if you have answered/chosen item [2, 3] in question 143, skip the following question*
*Note: if you have answered/chosen item [2, 3] in question 158, skip the following question*
*Note: if you have answered/chosen item [2, 3] in question 173, skip the following question*

**Q185: How many of the acute day unit staff are clinicians with mental health professional qualifications (e.g. nurses or social workers)?**

○ All of the staff ○ Most of the staff ○ Some of the staff ○ None of the staff ○ Unsure

*Note: if you have answered/chosen item [2, 3] in question 143, skip the following question*
*Note: if you have answered/chosen item [2, 3] in question 158, skip the following question*
*Note: if you have answered/chosen item [2, 3] in question 173, skip the following question*

**Q186: Is there any shared staffing between this acute day unit and any other local crisis service?**

○ Yes (please state below which teams and describe the arrangement briefly)
○ No
○ Unsure

*Note: if you have answered/chosen item [2, 3] in question 143, skip the following question*
*Note: if you have answered/chosen item [2, 3] in question 158, skip the following question*
*Note: if you have answered/chosen item [2, 3] in question 173, skip the following question*

**Q187: Is there any joint management at team-level between this acute day unit and any other local crisis service?**

○ Yes (please state below which teams and describe the arrangement briefly)
○ No
○ Unsure

*Note: if you have answered/chosen item [2, 3] in question 143, skip the following question*
*Note: if you have answered/chosen item [2, 3] in question 158, skip the following question*
*Note: if you have answered/chosen item [2, 3] in question 173, skip the following question*

**Q188: Are there any other acute day units in your area?**

◯ Yes ◯ No ◯ Unsure

#### All other acute day units

*Note: if you have answered/chosen item [2, 3] in question 143, skip the following question*
*Note: if you have answered/chosen item [2, 3] in question 158, skip the following question*
*Note: if you have answered/chosen item [2, 3] in question 173, skip the following question*
*Note: if you have answered/chosen item [2, 3] in question 188, skip the following question*

**Q189: Please list all other acute day units:**

#### Crisis family placements

*A service where local families offer short-term crisis foster placements in their family home for people in mental health crisis, supported by local crisis services eg the CRHTT – please do not tell us about respite family placements here – only placements of people in mental health crisis*

**Q190: Is there a crisis family placement scheme in your area?**

◯ Yes ◯ No ◯ Unsure

#### Crisis Family Placements (cont.)

*Note: if you have answered/chosen item [2, 3] in question 190, skip the following question*

**Q191: What is the name of the scheme?**

*Note: if you have answered/chosen item [2, 3] in question 190, skip the following question*

**Q192: What type of organisation provides the scheme?**

◯ NHS ◯ Local authority ◯ Voluntary sector ◯ Unsure

*Note: if you have answered/chosen item [2, 3] in question 190, skip the following question*

**Q193: Referral pathways (please select all that apply):**

- ☐ The CRHTT and/or crisis assessment service can refer to this service
- ☐ The CRHTT and/or crisis assessment service have exclusive referral rights
- ☐ People in crisis can self-refer to this service
- ☐ Unsure

Please provide any further information below which will help us understand the referral pathways into the crisis family placement service.

*Note: if you have answered/chosen item [2, 3] in question 190, skip the following question*

**Q194: What is the lower age limit for people who can use the family placement scheme? Please enter 0 if no minimum or 999 if unsure.**

*Note: if you have answered/chosen item [2, 3] in question 190, skip the following question*

**Q195: What is the upper age limit for people who can use the family placement scheme? Please enter 888 if no minimum or 999 if unsure.**

*Note: if you have answered/chosen item [2, 3] in question 190, skip the following question*

**Q196: Is this family placement scheme for any other specific patient group (e.g. specific diagnosis or demographic group, risk thresholds etc.)?**

- ○ Yes (please state which groups below)
- ○ No
- ○ Unsure

*Note: if you have answered/chosen item [2, 3] in question 190, skip the following question*

**Q197: Can the CRHTT or crisis assessment service access a place at this service for a patient when needed?**

- ◯ Always
- ◯ Usually
- ◯ Sometimes
- ◯ Rarely
- ◯ Unsure

*Note: if you have answered/chosen item [2, 3] in question 190, skip the following question*

**Q198: How quickly can admission to this service typically be arranged following initial referral?**

- ◯ In less than four hours
- ◯ Within 24 hours
- ◯ Within 72 hours
- ◯ Within one week
- ◯ Longer than one week
- ◯ Unsure

*Note: if you have answered/chosen item [2, 3] in question 190, skip the following question*

**Q199: What is the maximum length of stay with a family? Please enter 999 if unsure.**

[ ] days

*Note: if you have answered/chosen item [2, 3] in question 190, skip the following question*

**Q200: What is a typical length of stay with a family? Please enter 999 if unsure.**

[ ] days

*Note: if you have answered/chosen item [2, 3] in question 190, skip the following question*

**Q201: Are there any NHS mental health services that provide support for families offering the crisis family placements?**

- ◯ Yes (please state which services below)
- ◯ No
- ◯ Unsure

#### Crisis café / crisis drop-in services

**Q202: Is there a crisis café or crisis drop-in service in your local area?**

**This type of service might otherwise be referred to as a “Haven” or “sanctuary”. Here, we are asking about services which provides an out of hours assessment and immediate support for people in mental health crisis, as an alternative to attending Accident and Emergency.**

◯ Yes ◯ No ◯ Unsure

#### Crisis café / crisis drop-in service one

*Note: if you have answered/chosen item [2, 3] in question 202, skip the following question*

**Q203: What is the name of this service?**

*Note: if you have answered/chosen item [2, 3] in question 202, skip the following question*

**Q204: What type of organisation provides the service?**

◯ NHS ◯ Local authority ◯ Voluntary sector ◯ Unsure

*Note: if you have answered/chosen item [2, 3] in question 202, skip the following question*

**Q205: What are the opening hours?**

◯ 24hr
◯ 24hr Monday to Friday and 9am – 5pm Saturday and Sunday
◯ 24hr Monday to Friday
◯ 9am – 5pm Monday to Sunday
◯ 9am – 5pm Monday to Friday
◯ Other (please detail below)
◯ Unsure

*Note: if you have answered/chosen item [2, 3] in question 202, skip the following question*

**Q206: What is the lower age limit for people who can use this crisis café? Please enter 0 if no minimum or 999 if unsure.**

*Note: if you have answered/chosen item [2, 3] in question 202, skip the following question*

**Q207: What is the upper age limit? Please enter 888 if no minimum or 999 if unsure.**

*Note: if you have answered/chosen item [2, 3] in question 202, skip the following question*

**Q208: Is this crisis café for any other specific patient group (e.g. specific diagnosis or demographic group, risk thresholds etc.)?**

- ◯ Yes (please state which groups below)
- ◯ No
- ◯ Unsure

*Note: if you have answered/chosen item [2, 3] in question 202, skip the following question*

**Q209: Is this crisis café service user-led?**

- ◯ Yes
- ◯ No
- ◯ Unsure

*Note: if you have answered/chosen item [2, 3] in question 202, skip the following question*

**Q210: How many of the crisis café staff are clinicians with mental health professional qualifications (e.g. nurses or social workers)?**

- ◯ All of the staff
- ◯ Most of the staff
- ◯ Some of the staff
- ◯ None of the staff
- ◯ Unsure

*Note: if you have answered/chosen item [2, 3] in question 202, skip the following question*

**Q211: Is there any shared staffing between this crisis café and any other local crisis service?**

- ◯ Yes (please state below which teams and describe the arrangement briefly)
- ◯ No
- ◯ Unsure

*Note: if you have answered/chosen item [2, 3] in question 202, skip the following question*

**Q212: Is there any joint management at team-level between this crisis café and any other local crisis service?**

- ◯ Yes (please state below which teams and describe the arrangement briefly)
- ◯ No
- ◯ Unsure

*Note: if you have answered/chosen item [2, 3] in question 202, skip the following question*

**Q213: Can people self-refer to this crisis café?**

- ◯ Yes, they can drop-in without any appointment
- ◯ Yes, they can self-refer by phone but can only attend the service once they are booked in
- ◯ No, a referral from other health services is needed
- ◯ Unsure

Please provide any further information below which will help us understand the referral pathways into the crisis café.

*Note: if you have answered/chosen item [2, 3] in question 202, skip the following question*

**Q214: Can the CRHTT / crisis assessment team make referrals to this service?**

◯ Yes ◯ No ◯ Unsure

*Note: if you have answered/chosen item [2, 3] in question 202, skip the following question*

**Q215: Can this crisis café refer current users of the service to the CRHTT?**

◯ Yes ◯ No ◯ Unsure

*Note: if you have answered/chosen item [2, 3] in question 202, skip the following question*

**Q216: Can current users of this crisis café self-refer to the CRHTT?**

◯ Yes ◯ No ◯ Unsure

*Note: if you have answered/chosen item [2, 3] in question 202, skip the following question*

**Q217: Are there any other crisis cafés or crisis drop-in services in your area?**

◯ Yes ◯ No ◯ Unsure

#### Crisis café / crisis drop-in services two

*Note: if you have answered/chosen item [2, 3] in question 202, skip the following question*
*Note: if you have answered/chosen item [2, 3] in question 217, skip the following question*

**Q218: What is the name of this service?**

*Note: if you have answered/chosen item [2, 3] in question 202, skip the following question*
*Note: if you have answered/chosen item [2, 3] in question 217, skip the following question*

**Q219: What type of organisation provides the service?**

◯ NHS ◯ Local authority ◯ Voluntary sector ◯ Unsure

*Note: if you have answered/chosen item [2, 3] in question 202, skip the following question*
*Note: if you have answered/chosen item [2, 3] in question 217, skip the following question*

**Q220: What are the opening hours?**

◯ 24hr
◯ 24hr Monday to Friday and 9am – 5pm Saturday and Sunday
◯ 24hr Monday to Friday
◯ 9am – 5pm Monday to Sunday
◯ 9am – 5pm Monday to Friday
◯ Other (please detail below)
◯ Unsure

*Note: if you have answered/chosen item [2, 3] in question 202, skip the following question*
*Note: if you have answered/chosen item [2, 3] in question 217, skip the following question*

**Q221: What is the lower age limit for people who can use this crisis café? Please enter 0 if no minimum or 999 if unsure.**

*Note: if you have answered/chosen item [2, 3] in question 202, skip the following question*
*Note: if you have answered/chosen item [2, 3] in question 217, skip the following question*

**Q222: What is the upper age limit? Please enter 888 if no minimum or 999 if unsure.**

*Note: if you have answered/chosen item [2, 3] in question 202, skip the following question*
*Note: if you have answered/chosen item [2, 3] in question 217, skip the following question*

**Q223: Is this crisis café for any other specific patient group (e.g. specific diagnosis or demographic group, risk thresholds etc.)?**

- ◯ Yes (please state which groups below)
- ◯ No
- ◯ Unsure

*Note: if you have answered/chosen item [2, 3] in question 202, skip the following question*
*Note: if you have answered/chosen item [2, 3] in question 217, skip the following question*

**Q224: Is this crisis café service user-led?**

◯ Yes ◯ No ◯ Unsure

*Note: if you have answered/chosen item [2, 3] in question 202, skip the following question*
*Note: if you have answered/chosen item [2, 3] in question 217, skip the following question*

**Q225: How many of the crisis café staff are clinicians with mental health professional qualifications (e.g. nurses or social workers)?**

○ All of the staff ○ Most of the staff ○ Some of the staff ○ None of the staff ○ Unsure

*Note: if you have answered/chosen item [2, 3] in question 202, skip the following question*
*Note: if you have answered/chosen item [2, 3] in question 217, skip the following question*

**Q226: Is there any shared staffing between this crisis café and any other local crisis service?**

○ Yes (please state below which teams and describe the arrangement briefly)
○ No
○ Unsure

*Note: if you have answered/chosen item [2, 3] in question 202, skip the following question*
*Note: if you have answered/chosen item [2, 3] in question 217, skip the following question*

**Q227: Is there any joint management at team-level between this crisis café and any other local crisis service?**

○ Yes (please state below which teams and describe the arrangement briefly)
○ No
○ Unsure

*Note: if you have answered/chosen item [2, 3] in question 202, skip the following question*
*Note: if you have answered/chosen item [2, 3] in question 217, skip the following question*

**Q228: Can people self-refer to this crisis café?**

○ Yes, they can drop-in without any appointment
○ Yes, they can self-refer by phone but can only attend the service once they are booked in
○ No, a referral from other health services is needed
○ Unsure

Please provide any further information below which will help us understand the referral pathways into the crisis café.

*Note: if you have answered/chosen item [2, 3] in question 202, skip the following question*
*Note: if you have answered/chosen item [2, 3] in question 217, skip the following question*

**Q229: Can the CRHTT / crisis assessment team make referrals to this service?**

◯ Yes ◯ No ◯ Unsure

*Note: if you have answered/chosen item [2, 3] in question 202, skip the following question*
*Note: if you have answered/chosen item [2, 3] in question 217, skip the following question*

**Q230: Can this crisis café refer current users of the service to the CRHTT?**

◯ Yes ◯ No ◯ Unsure

*Note: if you have answered/chosen item [2, 3] in question 202, skip the following question*
*Note: if you have answered/chosen item [2, 3] in question 217, skip the following question*

**Q231: Can current users of this crisis café self-refer to the CRHTT?**

◯ Yes ◯ No ◯ Unsure

*Note: if you have answered/chosen item [2, 3] in question 202, skip the following question*
*Note: if you have answered/chosen item [2, 3] in question 217, skip the following question*

**Q232: Are there any other crisis cafés or crisis drop-in services in your area?**

◯ Yes ◯ No ◯ Unsure

#### Crisis café / crisis drop-in services three

*Note: if you have answered/chosen item [2, 3] in question 202, skip the following question*
*Note: if you have answered/chosen item [2, 3] in question 217, skip the following question*
*Note: if you have answered/chosen item [2, 3] in question 232, skip the following question*

**Q233: What is the name of this service?**

*Note: if you have answered/chosen item [2, 3] in question 202, skip the following question*
*Note: if you have answered/chosen item [2, 3] in question 217, skip the following question*
*Note: if you have answered/chosen item [2, 3] in question 232, skip the following question*

**Q234: What type of organisation provides the service?**

○ NHS ○ Local authority ○ Voluntary sector ○ Unsure

*Note: if you have answered/chosen item [2, 3] in question 202, skip the following question*
*Note: if you have answered/chosen item [2, 3] in question 217, skip the following question*
*Note: if you have answered/chosen item [2, 3] in question 232, skip the following question*

**Q235: What are the opening hours?**

○ 24hr
○ 24hr Monday to Friday and 9am – 5pm Saturday and Sunday
○ 24hr Monday to Friday
○ 9am – 5pm Monday to Sunday
○ 9am – 5pm Monday to Friday
○ Other (please detail below)
○ Unsure

*Note: if you have answered/chosen item [2, 3] in question 202, skip the following question*
*Note: if you have answered/chosen item [2, 3] in question 217, skip the following question*
*Note: if you have answered/chosen item [2, 3] in question 232, skip the following question*

**Q236: What is the lower age limit for people who can use this crisis café? Please enter 0 if no minimum or 999 if unsure.**

*Note: if you have answered/chosen item [2, 3] in question 202, skip the following question*
*Note: if you have answered/chosen item [2, 3] in question 217, skip the following question*
*Note: if you have answered/chosen item [2, 3] in question 232, skip the following question*

**Q237: What is the upper age limit? Please enter 888 if no minimum or 999 if unsure.**

*Note: if you have answered/chosen item [2, 3] in question 202, skip the following question*
*Note: if you have answered/chosen item [2, 3] in question 217, skip the following question*
*Note: if you have answered/chosen item [2, 3] in question 232, skip the following question*

**Q238: Is this crisis café for any other specific patient group (e.g. specific diagnosis or demographic group, risk thresholds etc.)?**

○ Yes (please state which groups below) ○ No
○ Unsure

*Note: if you have answered/chosen item [2, 3] in question 202, skip the following question*
*Note: if you have answered/chosen item [2, 3] in question 217, skip the following question*
*Note: if you have answered/chosen item [2, 3] in question 232, skip the following question*

**Q239: Is this crisis café service user-led?**

◯ Yes ◯ No ◯ Unsure

*Note: if you have answered/chosen item [2, 3] in question 202, skip the following question*
*Note: if you have answered/chosen item [2, 3] in question 217, skip the following question*
*Note: if you have answered/chosen item [2, 3] in question 232, skip the following question*

**Q240: How many of the crisis café staff are clinicians with mental health professional qualifications (e.g. nurses or social workers)?**

◯ All of the staff ◯ Most of the staff ◯ Some of the staff ◯ None of the staff ◯ Unsure

*Note: if you have answered/chosen item [2, 3] in question 202, skip the following question*
*Note: if you have answered/chosen item [2, 3] in question 217, skip the following question*
*Note: if you have answered/chosen item [2, 3] in question 232, skip the following question*

**Q241: Is there any shared staffing between this crisis café and any other local crisis service?**

◯ Yes (please state below which teams and describe the arrangement briefly)
◯ No
◯ Unsure

*Note: if you have answered/chosen item [2, 3] in question 202, skip the following question*
*Note: if you have answered/chosen item [2, 3] in question 217, skip the following question*
*Note: if you have answered/chosen item [2, 3] in question 232, skip the following question*

**Q242: Is there any joint management at team-level between this crisis café and any other local crisis service?**

◯ Yes (please state below which teams and describe the arrangement briefly)
◯ No
◯ Unsure

*Note: if you have answered/chosen item [2, 3] in question 202, skip the following question*
*Note: if you have answered/chosen item [2, 3] in question 217, skip the following question*
*Note: if you have answered/chosen item [2, 3] in question 232, skip the following question*

**Q243: Can people self-refer to this crisis café?**

- ◯ Yes, they can drop-in without any appointment
- ◯ Yes, they can self-refer by phone but can only attend the service once they are booked in
- ◯ No, a referral from other health services is needed
- ◯ Unsure

Please provide any further information below which will help us understand the referral pathways into the crisis café.

*Note: if you have answered/chosen item [2, 3] in question 202, skip the following question*
*Note: if you have answered/chosen item [2, 3] in question 217, skip the following question*
*Note: if you have answered/chosen item [2, 3] in question 232, skip the following question*

**Q244: Can the CRHTT / crisis assessment team make referrals to this service?**

◯ Yes ◯ No ◯ Unsure

*Note: if you have answered/chosen item [2, 3] in question 202, skip the following question*
*Note: if you have answered/chosen item [2, 3] in question 217, skip the following question*
*Note: if you have answered/chosen item [2, 3] in question 232, skip the following question*

**Q245: Can this crisis café refer current users of the service to the CRHTT?**

◯ Yes ◯ No ◯ Unsure

*Note: if you have answered/chosen item [2, 3] in question 202, skip the following question*
*Note: if you have answered/chosen item [2, 3] in question 217, skip the following question*
*Note: if you have answered/chosen item [2, 3] in question 232, skip the following question*

**Q246: Can current users of this crisis café self-refer to the CRHTT?**

◯ Yes ◯ No ◯ Unsure

*Note: if you have answered/chosen item [2, 3] in question 202, skip the following question*
*Note: if you have answered/chosen item [2, 3] in question 217, skip the following question*
*Note: if you have answered/chosen item [2, 3] in question 232, skip the following question*

**Q247: Are there any other crisis cafés or crisis drop-in services in your area?**

◯ Yes ◯ No ◯ Unsure

#### Other crisis café / crisis drop-in services

*Note: if you have answered/chosen item [2, 3] in question 202, skip the following question*
*Note: if you have answered/chosen item [2, 3] in question 217, skip the following question*
*Note: if you have answered/chosen item [2, 3] in question 232, skip the following question*
*Note: if you have answered/chosen item [2, 3] in question 247, skip the following question*

**Q248: Please list all other crisis cafés / crisis drop-in services in your area:**

#### Police street triage service

**Q249: Is there a local street triage service where mental health staff work jointly with the police?**

**A team where mental health staff work jointly with the police services to arrange appropriate help for people with mental health crisis who come to the attention of the police, and provide alternatives to using s.136 where possible.**

◯ Yes ◯ No ◯ Unsure

#### Police street triage service (cont.)

*Note: if you have answered/chosen item [2, 3] in question 249, skip the following question*

**Q250: What is the name of this service?**

*Note: if you have answered/chosen item [2, 3] in question 249, skip the following question*

**Q251: What are the opening hours?**

- ◯ 24hr
- ◯ 24hr Monday to Friday and 9am – 5pm Saturday and Sunday
- ◯ 24hr Monday to Friday
- ◯ 9am – 5pm Monday to Sunday
- ◯ 9am – 5pm Monday to Friday
- ◯ Other (please detail below)
- ◯ Unsure

*Note: if you have answered/chosen item [2, 3] in question 249, skip the following question*

**Q252: What is the model of care?**

- ◯ Mental health worker based in police call centre – phone advice only
- ◯ Based in police call centre – can go out with police
- ◯ Mobile unit with mental health worker and police
- ◯ Other (please describe below)
- ◯ Unsure

*Note: if you have answered/chosen item [2, 3] in question 249, skip the following question*

**Q253: Is there any shared staffing between the street triage service and any other mental health crisis service?**

- ◯ Yes (please state below which teams and describe the arrangement briefly)
- ◯ No
- ◯ Unsure

*Note: if you have answered/chosen item [2, 3] in question 249, skip the following question*

**Q254: Is there any joint management at team-level between the street triage service and any other local mental health crisis service?**

- ◯ Yes (please state below which teams and describe the arrangement briefly)
- ◯ No
- ◯ Unsure

#### Police street triage service (cont.)

*You have told us there is no separate police street triage service in your local area. Now please tell us...*

*Note: if you have answered/chosen item [1] in question 249, skip the following question*

**Q255: Does the CRHTT and/or crisis assessment service attend assessments with the police in public places?**

◯ Yes ◯ No ◯ Unsure

*Note: if you have answered/chosen item [1] in question 249, skip the following question*

**Q256: Can the police bring people (not on s.136) to health premises for CRHTT or crisis assessment service assessment?**

◯ Yes ◯ No ◯ Unsure

#### Ambulance street triage service

**Q257: Is there a local street triage service where mental health staff work jointly with ambulance service staff?**

**A team where mental health staff work jointly with the ambulance services to arrange appropriate help for people with mental health crisis who come to the attention of the police, and provide alternatives to using s.136 where possible.**

◯ Yes ◯ No ◯ Unsure

#### Ambulance street triage service (cont.)

*Note: if you have answered/chosen item [2, 3] in question 257, skip the following question*

**Q258: What is the name of this service?**

*Note: if you have answered/chosen item [2, 3] in question 257, skip the following question*

**Q259: What are the opening hours?**

- ◯ 24hr
- ◯ 24hr Monday to Friday and 9am – 5pm Saturday and Sunday
- ◯ 24hr Monday to Friday
- ◯ 9am – 5pm Monday to Sunday
- ◯ 9am – 5pm Monday to Friday
- ◯ Other (please detail below)
- ◯ Unsure

*Note: if you have answered/chosen item [2, 3] in question 257, skip the following question*

**Q260: What is the model of care?**

- ◯ Mental health worker based in ambulance service call centre – phone advice only
- ◯ Based in ambulance service call centre – can go out with police
- ◯ Mobile unit with mental health worker and ambulance staff
- ◯ Other (please describe below)
- ◯ Unsure

*Note: if you have answered/chosen item [2, 3] in question 257, skip the following question*

**Q261: Is there any shared staffing between the street triage service and any other mental health crisis service?**

- ◯ Yes (please state below which teams and describe the arrangement briefly)
- ◯ No
- ◯ Unsure

*Note: if you have answered/chosen item [2, 3] in question 257, skip the following question*

**Q262: Is there any joint management at team-level between the street triage service and any other local mental health crisis service?**

- ◯ Yes (please state below which teams and describe the arrangement briefly)
- ◯ No
- ◯ Unsure

#### Ambulance street triage service (cont.)

*You have told us there is no separate ambulance street triage service in your local area. Now please tell us...*

*Note: if you have answered/chosen item [1] in question 257, skip the following question*

**Q263: Does the CRHTT and/or crisis assessment service attend assessments with the ambulance staff in public places?**

◯ Yes ◯ No ◯ Unsure

*Note: if you have answered/chosen item [1] in question 257, skip the following question*

**Q264: Can the ambulance bring people (not on s.136) to health premises for CRHTT or crisis assessment service assessment?**

◯ Yes ◯ No ◯ Unsure

#### Any other local crisis services

**Q265: Are there any other local crisis services which the CRHTT or crisis assessment service often works with?**

◯ Yes ◯ No ◯ Unsure

#### Any other local crisis services (cont.)

*Note: if you have answered/chosen item [2, 3] in question 265, skip the following question*

**Q266: Please list the names of all other local crisis services, and provide a brief description of each:**

#### Outcomes monitoring

**Q267: Does your CRHTT service routinely use Patient-Reported Outcome Measures (PROMs)?**

**PROMs are self-report measures completed by patients of outcomes relating to clinical recovery (e.g. symptoms or social functioning).**

◯ Yes ◯ No ◯ Unsure

**Q268: Does your CRHTT service routinely use Patient-Reported Experience Measures (PREMs)?**

**PREMs are self-report measures completed by patients relating to satisfaction with care or quality of life.**

◯ Yes ◯ No ◯ Unsure

**Q269: Does your CRHTT service routinely use Clinician-Reported Outcome Measures (CROMs)?**

**CROMs are structured measures of symptoms or functioning completed by staff based on their knowledge of the patient and clinical judgement.**

◯ Yes ◯ No ◯ Unsure

### Outcomes monitoring: PROMs

*You have told us your CRHTT regularly use PROMs. Now please tell us...*

*Note: if you have answered/chosen item [2, 3] in question 267, skip the following question*

**Q270: Which outcome measurement tools are used? Please list. Please enter 999 if unsure.**

*Note: if you have answered/chosen item [2, 3] in question 267, skip the following question*

**Q271: What is the method of data collection (e.g. phone interview, postal questionnaire, online survey)? Please state. Enter 999 if unknown.**

*Note: if you have answered/chosen item [2, 3] in question 267, skip the following question*

**Q272: Do you collect data at:**

- ◯ Discharge only
- ◯ Admission and discharge
- ◯ Other (please describe below)
- ◯ Unsure

*Note: if you have answered/chosen item [2, 3] in question 267, skip the following question*

**Q273: What are the response rates?**

**If you collect data at admission and discharge from the service, please tell us the % of patients from whom you obtain data at both time points. Enter 999 if unknown.**

%

*Note: if you have answered/chosen item [2, 3] in question 267, skip the following question*

**Q274: Is this response rate an estimate or based on data?**

◯ Estimate ◯ Based on data ◯ Not applicable ◯ Unsure

*Note: if you have answered/chosen item [2, 3] in question 267, skip the following question*

**Q275: Are these outcome measures used in other crisis services within the Trust?**

◯ Yes (please state which services below) ◯ No
◯ Unsure

### Outcomes monitoring: PREMs

*You have told us your CRHTT regularly use PREMs. Now please tell us...*

*Note: if you have answered/chosen item [2, 3] in question 268, skip the following question*

**Q276: Which patient experience measurement tools are used? Please list. Please enter 999 if unsure.**

*Note: if you have answered/chosen item [2, 3] in question 268, skip the following question*

**Q277: What is the method of data collection (e.g. phone interview, postal questionnaire, online survey)? Please state. Enter 999 if unknown.**

*Note: if you have answered/chosen item [2, 3] in question 268, skip the following question*

**Q278: What are the response rates?**

**If you collect data at admission and discharge from the service, please tell us the % of patients from whom you obtain data at both time points. Enter 999 if unknown.**

[ ] %

*Note: if you have answered/chosen item [2, 3] in question 268, skip the following question*

**Q279: Is this response rate an estimate or based on data?**

○ Estimate ○ Based on data ○ Unsure

*Note: if you have answered/chosen item [2, 3] in question 268, skip the following question*

**Q280: Are these outcome measures used in other crisis services within the Trust?**

○ Yes (please state which services below) ○ No
○ Unsure

#### Outcomes monitoring: CROMs

*You have told us your CRHTT regularly use CROMs. Now please tell us...*

*Note: if you have answered/chosen item [2, 3] in question 269, skip the following question*

**Q281: Which outcome measurement tools are used? Please list. Please enter 999 if unsure.**

*Note: if you have answered/chosen item [2, 3] in question 269, skip the following question*

**Q282: Do you collect data at:**

○ Discharge only ○ Admission and discharge ○ Other (please describe below)
○ Unsure

*Note: if you have answered/chosen item [2, 3] in question 269, skip the following question*

**Q283: What are the response rates? Enter 999 if unknown.**

[ ] %

*Note: if you have answered/chosen item [2, 3] in question 269, skip the following question*

**Q284: Is this response rate an estimate or based on data?**

○ Estimate ○ Based on data ○ Unsure

*Note: if you have answered/chosen item [2, 3] in question 269, skip the following question*

**Q285: Are these outcome measures used in other crisis services within the Trust?**

○ Yes (please state which services below) ○ No
○ Unsure

#### Further information

**Q286: Is there an overall manager or leadership group for mental health crisis / acute services in your local area?**

○ Yes ○ No ○ Unsure

#### Further information (cont.)

*Note: if you have answered/chosen item [2, 3] in question 286, skip the following question*

**Q287: What is the manager's or group's name?**

*Note: if you have answered/chosen item [2, 3] in question 286, skip the following question*

**Q288: What is the geographical or administrative area for which they lead crisis services? Please enter 999 if unsure.**

*Note: if you have answered/chosen item [2, 3] in question 286, skip the following question*

**Q289: Do they manage all the services you have told us about in this survey?**

- ◯ Yes
- ◯ No (please state below which services they do manage)
- ◯ Unknown

#### Thank you

*Thank you for your help completing this survey. In order to obtain any information you are unsure about, or for accuracy checks on a sample of survey responses, we would like to contact another person from your NHS Trust with a good knowledge of local mental health crisis services (e.g. a local Acute Care Lead). Please could you provide the name and contact details for the most suitable person we could contact?*

**Q290: Name:**

**Q291: Job title:**

**Q292: Email address:**
